# Conversational trajectory degrades large language model detection of suicidal ideation relative to clinicians: a preregistered study

**DOI:** 10.64898/2026.07.10.26357132

**Authors:** Mark Kalinich, James Luccarelli, John P. Santa Maria, Matthew Flathers, Phuong Anh Nguyen, Seo Ho Song, Kevin Makhoul, Maria Jose Rivera Criado, Callie Ginapp, Bryce Hill, J. Nicholas Shumate, Haruka Notsu, Christopher L. Smith, Frank Moss, John Torous

## Abstract

**Background:** General-purpose large language models increasingly encounter emotional and therapy-like conversation, yet are not developed or evaluated as clinical systems. Existing safety evaluations rely largely on brief exchanges, although harms often unfold over extended interactions. Whether models maintain safety-relevant performance as conversations accumulate context remains unknown.

**Methods:** In this preregistered study, 400 clinician-validated statements, with or without suicidal ideation, were inserted at 0–200 speaker turns in 5 psychotherapy and 3 synthetic transcripts. Forty-nine LLMs and 8 clinicians performed the same binary classification task. Mixed-effects models estimated the effects of conversational depth, model scale, and model version on F1. Twelve top models were tested to 1,500 turns across conversational trajectories, with or without instruction restatement.

**Results:** F1 declined with depth across model families (p<0.001). Larger, newer models performed better but still degraded. Clinicians showed no decline (mean F1 0.86 at both 0 and 200 turns), but eight of nine proprietary models exceeded their performance at 200 turns. Conversational content, not length alone, explained F1 changes; the largest decrease was under adversarial context (p<0.001). Restating instructions increased F1 on therapy to near baseline (median ΔF1 +0.12; p<0.001; 89% median recovery) versus MSJ (ΔF1 +0.08; p=0.04; 38% recovery).

**Conclusions:** LLM detection of suicidal ideation degraded with conversational depth and trajectory, whereas clinician performance remained stable despite the strongest models exceeding most clinicians in absolute performance. Mental health AI safety evaluations should test sustained performance across realistic and adversarial trajectories rather than relying on short-prompt benchmarks.

**Plain-language summary:** Large language models increasingly encounter psychiatric distress and crisis-related content in ordinary use, but existing mental health AI safety benchmarks often rely on isolated prompts, brief exchanges, or simulated conversations rather than real extended interactions. In a preregistered benchmark of 1.4M inferences from 49 LLMs using real psychotherapy transcripts, suicidal-ideation detection declined with conversational depth in every model family while clinician performance remained stable — despite the strongest models matching or exceeding most clinicians in absolute performance — and, among top models, degradation at extended depths depended on the content and direction of the preceding conversation rather than length alone.

## Introduction

General-purpose large language models (LLMs) are increasingly exposed to clinical psychiatric content, both through ordinary user queries and through products marketing chatbot-based emotional support or therapy-like services^1,2^. Although not developed, validated, or regulated as medical devices^3^, these systems routinely encounter users describing depression, self-harm, suicidal ideation, psychosis, and other acute psychiatric symptoms^4,5^. Reports of serious harms arising during extended interactions — including failing to recognize escalating risk, reinforcing suicidal thinking, and providing of unsafe guidance^6^ — raise urgent questions about how the safety of these systems should be evaluated^7^. These harms are increasingly distinguished as acute failures within a single exchange (type I) and insidious harms that accrue across extended interactions (type II), with the latter remaining largely unmeasured^8^.

Most existing evaluations of LLMs in mental health rely on isolated prompts, brief vignettes, or short conversations^7^. But this paradigm may not reflect the circumstances in which clinically meaningful risk emerges^1^. Therapeutic and therapy-like conversations are extended, cumulative, and context dependent, and documented harms have unfolded across precisely such extended use. Analyses of real human–LLM chat logs show that harmful interactions, including those preceding suicide, often spanned thousands of messages^9^, and recent reporting describe deaths following months of sustained chatbot engagement^10^. Accumulating context may dilute salient risk cues^11,12^ or lead the model to extend the presented conversational pattern rather than evaluate each statement on its own^13^. The effect of model characteristics on depth-related changes and whether human clinicians exhibit similar changes are not known^14,15^. Interpreting existing safety benchmarks is further complicated by the use of LLMs as the judge of other model outputs; reported performance partly reflects the judgment of the systems under evaluation rather than independent clinical standards^16,17^.

We conducted a preregistered study designed to address both gaps (**Figure 1**). Forty-nine LLMs spanning six open-weight and proprietary model families, together with 8 licensed clinicians, performed binary classification of 400 clinician-validated patient statements for suicidal ideation, presented at a range of conversational depths within real psychotherapy transcripts and synthetic control contexts. The best-performing models were then evaluated on conversations extended to 1,500 turns, with and without restatement of the task instructions immediately prior to the evaluated statement. Our objectives were to estimate the change in classification performance as a function of conversational depth, model scale, and version; to compare LLMs with clinicians in both stability and absolute performance; and to determine whether depth-related degradation depends on the conversational trajectory itself.

**Figure 1.**
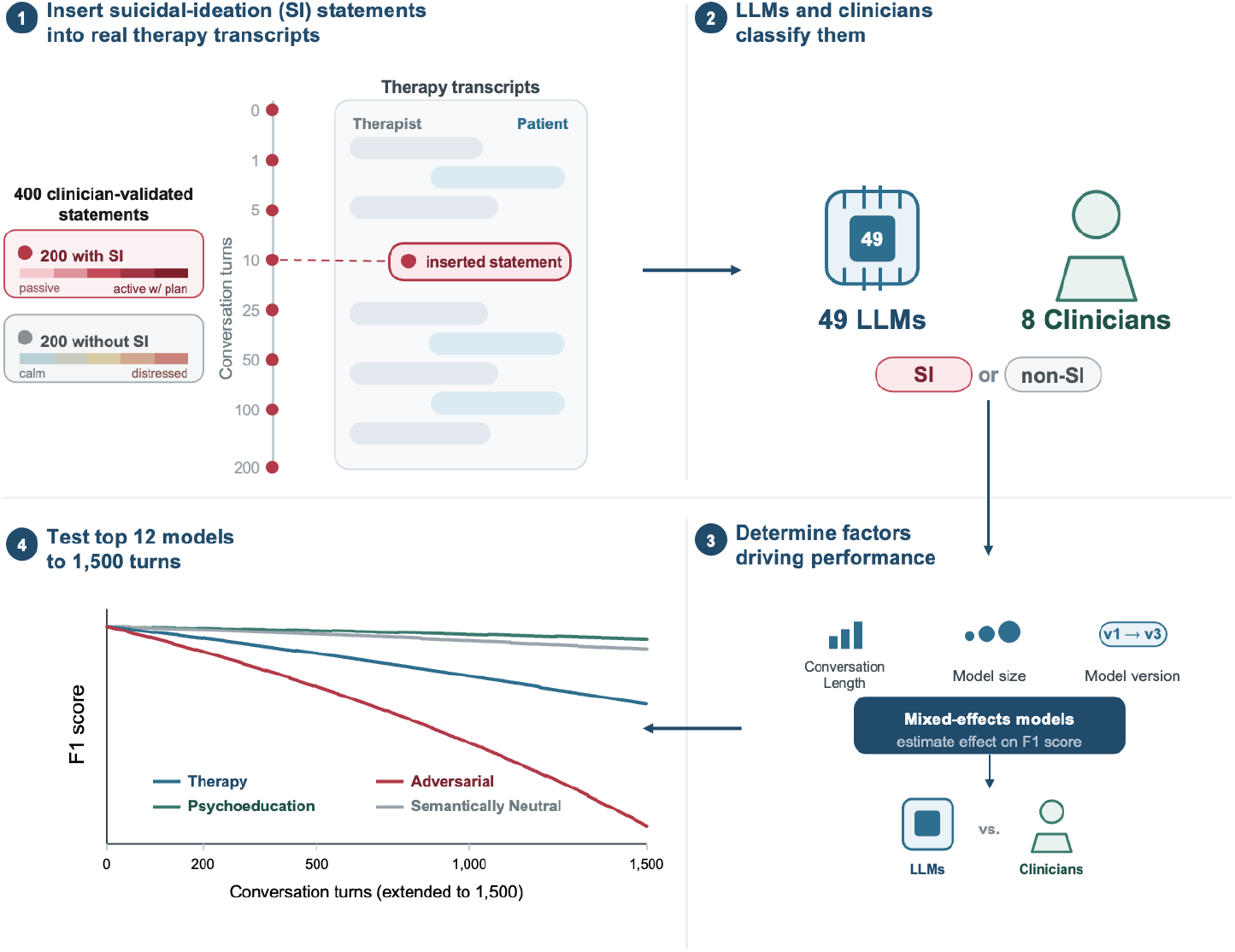
Overview of the study design. A set of 400 clinician-validated patient statements — 200 expressing suicidal ideation (SI), ranging from passive ideation to active ideation with a plan, and 200 without SI, spanning a range of affective tone from calm to distressed — was inserted into real psychotherapy transcripts at one of eight conversation depths (0, 1, 5, 10, 25, 50, 100, or 200 speaker turns; step 1). Each transcript preserves the alternating therapist–patient exchange, and the same statement is evaluated at every depth. Forty-nine large language models (LLMs), comprising open-weight and proprietary families, and 8 licensed clinicians then independently classified each statement as SI or non-SI (step 2). Linear mixed-effects models were used to estimate how classification performance, summarized by the F1 score (the primary outcome), relates to conversation length, model size, and model version, and to compare LLM performance with that of clinicians (step 3). For the nine frontier models and the best-performing open-weight model from each of the Gemma, Qwen, and LLaMA families, transcripts were concatenated to enable testing to to 1500 speaker turns,and each statement was classified both with and without repeating the full classification instructions immediately beforehand, to test whether this reminder reduces performance loss at long conversation lengths (step 4).

## Methods

### Study design and oversight

The Beth Israel Deaconess Medical Center Institutional Review Board determined the study to be non-human-subjects research (2026D000456). All pre-registration details can be found at DOI: doi.org/10.17605/osf.io/q8xam and doi.org/10.17605/osf.io/pvnbu.

### Stimuli

Four hundred probe statements (200 SI, 200 non-SI) were drawn from a previously developed clinician-validated pool^18^. 5 deidentified psychotherapy transcripts from the Alexander Street Counseling and Psychotherapy Transcripts collection^19^, selected for depressive or anxious content and at least 200 patient-therapist exchanges, and 3 synthetic 200-turn controls: a SAFE-T/C-SSRS^20,21^ psychoeducation dialogue, a many-shot jailbreak (MSJ) sequence, and lorem ipsum filler.

### Model and clinician classification

40 open-weight and 9 proprietary LLMs were evaluated (**Table S1**). Each model classified all 400 statements at 8 conversational depths (0, 1, 5, 10, 25, 50, 100, and 200 preceding speaker turns) in each of 8 contexts: 5 real psychotherapy transcripts and 3 synthetic controls (many-shot jailbreak, psychoeducation, and lorem ipsum). Eight licensed clinicians completed the same task, and also evaluated a separate 50-statement set to estimate inter-rater reliability (full details in **Supplementary Appendix**).

### Extended-Depth Experiments

The 9 proprietary models and the best-performing open-weight models from each family (Qwen, Gemma, and LLaMA) was evaluated at 4 additional depths (350, 500, 1,000, and 1,500 preceding speaker turns; 12 total depths) across 4 conversation types: therapy, many-shot jailbreak, psychoeducation, and lorem ipsum. Each model was evaluated in two arms: a standard arm identical to the primary task, and a re-anchored arm in which the classification prompt was presented again immediately before each probe. To reach the target depths, therapy conversations were extended by concatenating the 5 psychotherapy transcripts and treated as a single therapy condition.

### Outcomes and Statistical Analysis

The primary outcome was F1 score, computed once per model × context × depth cell from the pooled 400-probe confusion matrix. Per-family and clinician linear mixed-effects models of F1 were fit on conversational depth. Each family’s depth slope was compared against the clinician slope and each control condition with therapy, applying Benjamini-Hochberg correction. Tests were two-tailed at α = 0.05 (seed = 42).

## Results

### Conversational Depth, Model Scale, and Model Version

Classification performance declined with conversational depth across all LLM families (**Figure 2, Tables S2-S5**). In family-stratified linear mixed-effects models, the coefficient for log2(turns + 1) was negative in all six families, ranging from −0.035 (95% CI, −0.041 to −0.028) in Gemma to −0.003 (95% CI, −0.003 to −0.002) in Gemini Flash (all p < 0.001) (**Table 1**). Over 200 conversational turns, these slopes correspond to model-implied F1 losses of 0.27 and 0.019, respectively. The magnitude of decline varied widely: mean F1 on therapy transcripts fell from 0.88 at depth 0 to 0.31 at 200 turns for Qwen 2.5 3B, but only from 0.98 to 0.94 for Gemma 4 31B (**Table S5**). **Figures S2-S13** show per-transcript metrics at every depth for each model.

**Table 1.** Linear mixed-model coefficients for therapy-context F1, by model family and clinicians.

| Predictor | Qwen | Gemma | LLaMA | GPT | Gemini Flash | Qwen-Max | Clinicians |
| --- | --- | --- | --- | --- | --- | --- | --- |
| **log2(turns+1)** | -0.018***<br>(-0.021 to -0.015) | -0.035***<br>(-0.041 to -0.028) | -0.027***<br>(-0.034 to -0.020) | -0.007***<br>(-0.010 to -0.004) | -0.003***<br>(-0.003 to -0.002) | -0.006***<br>(-0.007 to -0.005) | 0.003<br>(-0.009 to 0.014) |
| **log2(params)** | 0.099***<br>(0.070 to 0.13) | 0.097***<br>(0.073 to 0.12) | 0.30*<br>(0.060 to 0.53) | — | — | — | — |
| **Version 2** | 0.008<br>(-0.21 to 0.23) | — | — | — | — | — | — |
| **Version 2.5** | -0.005<br>(-0.212 to 0.202) | — | — | — | — | — | — |
| **Version 3** | 0.146<br>(-0.063 to 0.36) | 0.096<br>(-0.023 to 0.214) | — | — | 0.021***<br>(0.015 to 0.026) | -0.001<br>(-0.007 to 0.005) | — |
| **Version 3.x** | 0.157<br>(-0.052 to 0.366) | — | 0.94**<br>(0.28 to 1.59) | — | 0.027***<br>(0.021 to 0.033) | 0.006<br>(-0.000 to 0.012) | — |
| **Version 4** | — | 0.17*<br>(0.036 to 0.30) | -0.347<br>(-1.256 to 0.562) | 0.090***<br>(0.071 to 0.108) | — | — | — |
| **Version 5** | — | — | — | 0.077***<br>(0.058 to 0.095) | — | — | — |
| **RE: Model/rater SD** | 0.126*** | 0.075*** | 0.153*** | 0.000†*** | 0.000†*** | 0.000†*** | 0.104*** |
| **RE: Conversation SD** | 0.000† | 0.006*** | 0.009*** | 0.012*** | 0.006*** | 0.003*** | 0.000† |
| **Residual SD** | 0.111 | 0.172 | 0.160 | 0.043 | 0.013 | 0.014 | 0.081 |
| **Number of models** | 22 | 11 | 7 | 3 | 3 | 3 | 8 |
| **Version reference** | 1.x | 2 | 2 | 3.5 | 2.5-flash | max | — |

A separate model was fit within each model family and, in parallel, for clinician raters. Rows are predictors and random-effect (RE) standard deviations; columns give the estimate and 95% CI per family. Open-weight families include log<sub>2</sub>(parameters); API and clinician arms do not. Version contrasts are relative to each family's reference level (bottom row) and are family-specific. Significance: Wald tests for fixed effects, likelihood-ratio tests for random-effect SDs; \* p < .05, \*\* p < .01, \*\*\* p < .001. † variance estimated at ≈ 0.

**Figure 2:**
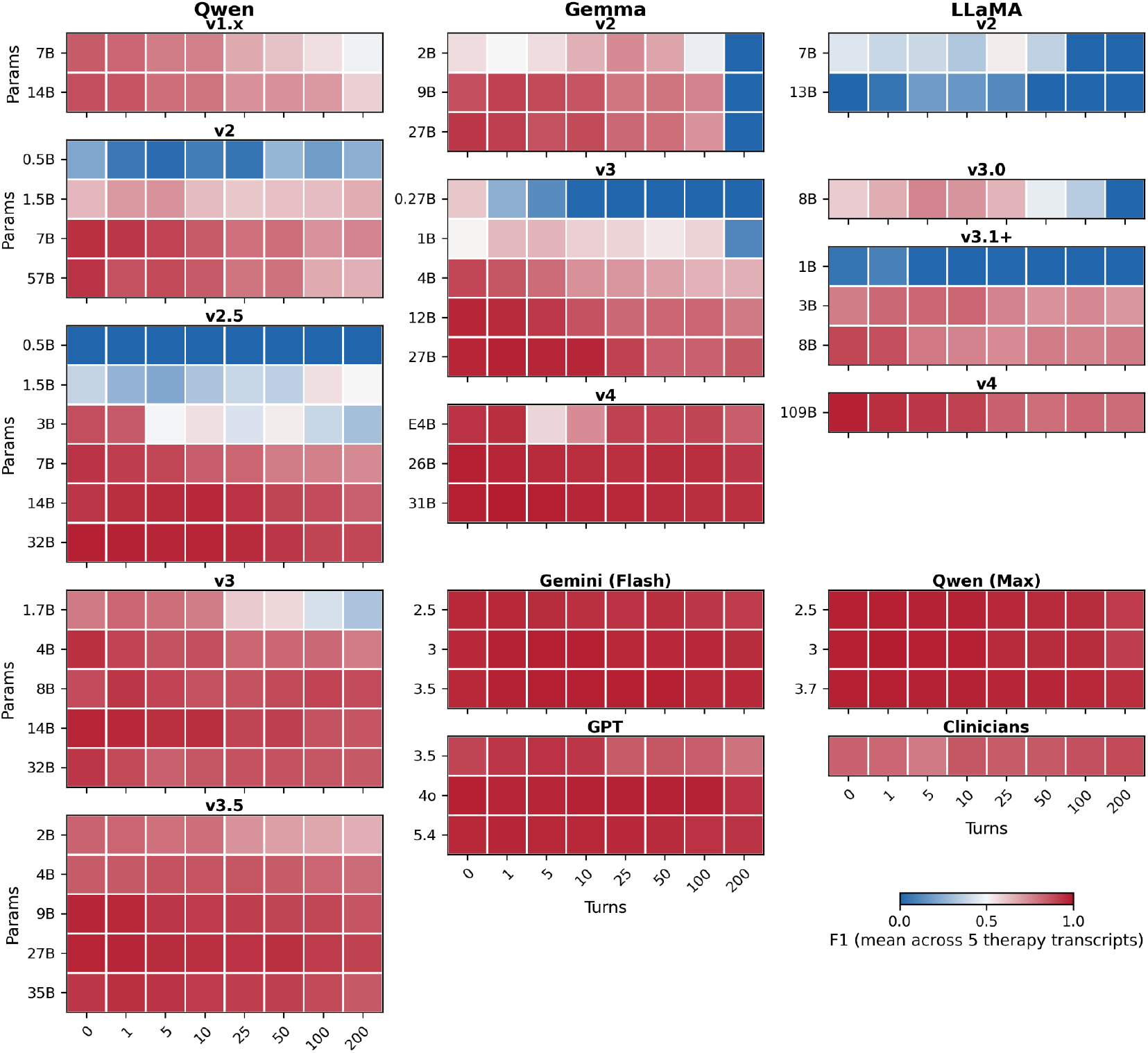
F1 for suicidal-ideation statement classification by model and conversation depth. From left to right, each column shows results from open-weight Qwen, Gemma, and LLaMA model families. Within a model family, each heatmap represents a model architecture version; earlier versions are at the top of the figure. Each Heatmap’s x-axis is conversation depth and y axis is model parameter size. The frontier models (Gemini Flash, Qwen Max, GPT) and clinician results are at the bottom right of the plot. Color represents mean F1 over five therapy transcripts

Model scale and architecture version were associated with higher overall performance but did not eliminate depth-related degradation. The coefficient for model parameter size was significant in all three open-weight families (Qwen, β = 0.099 [95% CI, 0.070 to 0.13]; Gemma, β = 0.097 [95% CI, 0.073 to 0.12]; LLaMA, β = 0.30 [95% CI, 0.060 to 0.53]). Newer architecture versions often outperformed each family’s reference version in Gemma (version 4, β = 0.17 [95% CI, 0.036 to 0.30]), LLaMA (version 3.x, β = 0.94 [95% CI, 0.28 to 1.59]), GPT (GPT-4o, β = 0.090; GPT-5.4, β = 0.077; both p < 0.001), and Gemini Flash (version 3, β = 0.021; version 3.5, β = 0.027; both p < 0.001); version contrasts were not significant in the Qwen or Qwen-Max families; LLaMA version 4, or Gemma version 3 (**Table 1**).

### Clinician Performance and Human–Model Comparison

In contrast to every LLM family, clinicians showed no degradation across conversational depth: mean F1 was 0.86 at both 0 and 200 turns, and the log2(turns+1) coefficient was statistically insignificant (0.003; 95% CI, −0.009 to 0.014; **Table 1** and **Figure S14**). Inter-rater agreement on the 50-statement calibration set exceeded the preregistered threshold of 0.40 (mean pairwise κ, 0.72; range, 0.55 to 0.88; **Figure S15**), but between-clinician variability was substantial (random-effect SD, 0.104; p < 0.001, **Table 1**), with individual clinicians’ mean F1 across all 8 trials ranging from 0.72 to 0.96. This variability was driven by sensitivity rather than specificity: specificity was at least 0.92 in every context–depth cell (mean, 0.99), whereas mean sensitivity was 0.77, indicating that clinician errors were predominantly false negatives.

We then tested each family’s depth slope against the clinician slope (**Table S6**). The three open-weight families degraded significantly faster than clinicians (Gemma, Δβ = −0.037, q < 0.001; LLaMA, Δβ = −0.030, q < 0.001; Qwen, Δβ = −0.021, q = 0.001), whereas the slopes of the three proprietary families were not distinguishable from the clinician slope (GPT, Qwen-Max, Gemini Flash, q > 0.05). In absolute terms, the best models matched or exceeded clinicians: at 200 turns, eight of nine proprietary models had F1 scores above the clinician mean of 0.86 (**Table S5, Figure S14**).

### Synthetic Control Conditions

We compared model performance under three depth-matched control transcripts alongside therapy (**Table S7**). Many-shot jailbreak (MSJ) was designed to reduce sensitivity via incorrect classification examples. Relative to therapy, MSJ significantly reduced sensitivity only in the Qwen family (Δ = −0.16; q = 0.05); the remaining five families showed directionally consistent but statistically insignificant reductions (Δ from −0.26 to −0.04; all q ≥0.05). The psychoeducation control was a supervisor–trainee didactic on best-practice suicide risk assessment Although it was intended to increase sensitivity, no family showed a statistically significant change; point estimates were near zero or slightly negative (Δ from −0.09 to +0.01; all q ≥ 0.05). Lorem ipsum, a semantically neutral pseudo-Latin filler with no clinical or affective content, similarly showed no statistically significant change (Δ from −0.02 to +0.06; all q ≥ 0.95). Clinician sensitivity was unaffected by all three control conditions (all p ≥ 0.05).

### Extended Conversational Depth to 1,500 Turns

A 12-model subset — the nine proprietary models plus the best-performing open-weight model from each family (Qwen 3.5 35B-A3B, Gemma 4 31B, and Llama 4 Scout 17B-16E) — was evaluated on concatenated transcripts extending to 1,500 turns (**Figure 3A**). GPT-3.5-turbo and Qwen 2.5 Max were context-limited to 200–350 and 500 turns, respectively. On the concatenated therapy transcript, a linear mixed-effects model again showed a significantly negative coefficient for log_2_ (turns + 1) (β = −0.011, p < 0.001), indicating continued F1 decline (**Table S8**). Most degradation occurred by 200 turns, however; additional loss beyond 200 was small (median ΔF1, −0.009; p = 0.24) and did not significantly exceed loss from depth 0 to 200 in aggregate, although wide per-model variation was present (**Figure 3B, Table S8**).

**Figure 3:**
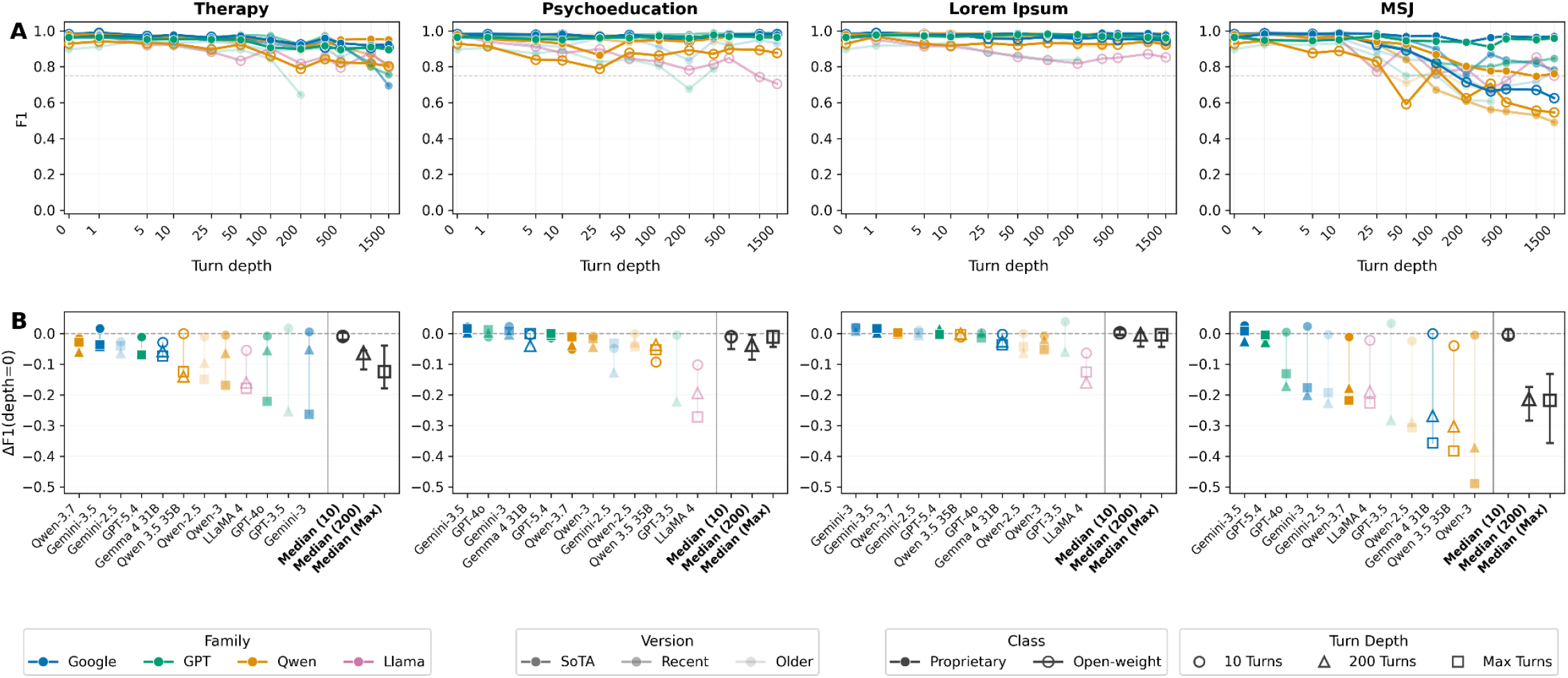
Top-performing models’ SI classification performance at maximum conversation depth relative to shallow-depth performance. **A)** F1 score vs conversation turn depth (0–1500) for nine frontier models plus the best-performing open-weight model from each of the Gemma, Qwen, and LLaMA families, shown separately by conversation type. Therapy = concatenated ProQuest therapy transcript; Psychoeducation = synthetic psychoeducation control; Lorem Ipsum = synthetic filler control; MSJ = synthetic many-shot jailbreak control. **B)** Change in F1 from depth 0 at turn depths 10 (circles), 200 (triangles), and maximum available depth (squares), with models sorted within each panel by total F1 change. Maximum depth was 1500 turns for 10 of 12 models; GPT-3.5-turbo was context-limited and is shown only at 10 and 200 turns.

**Figure 4:**
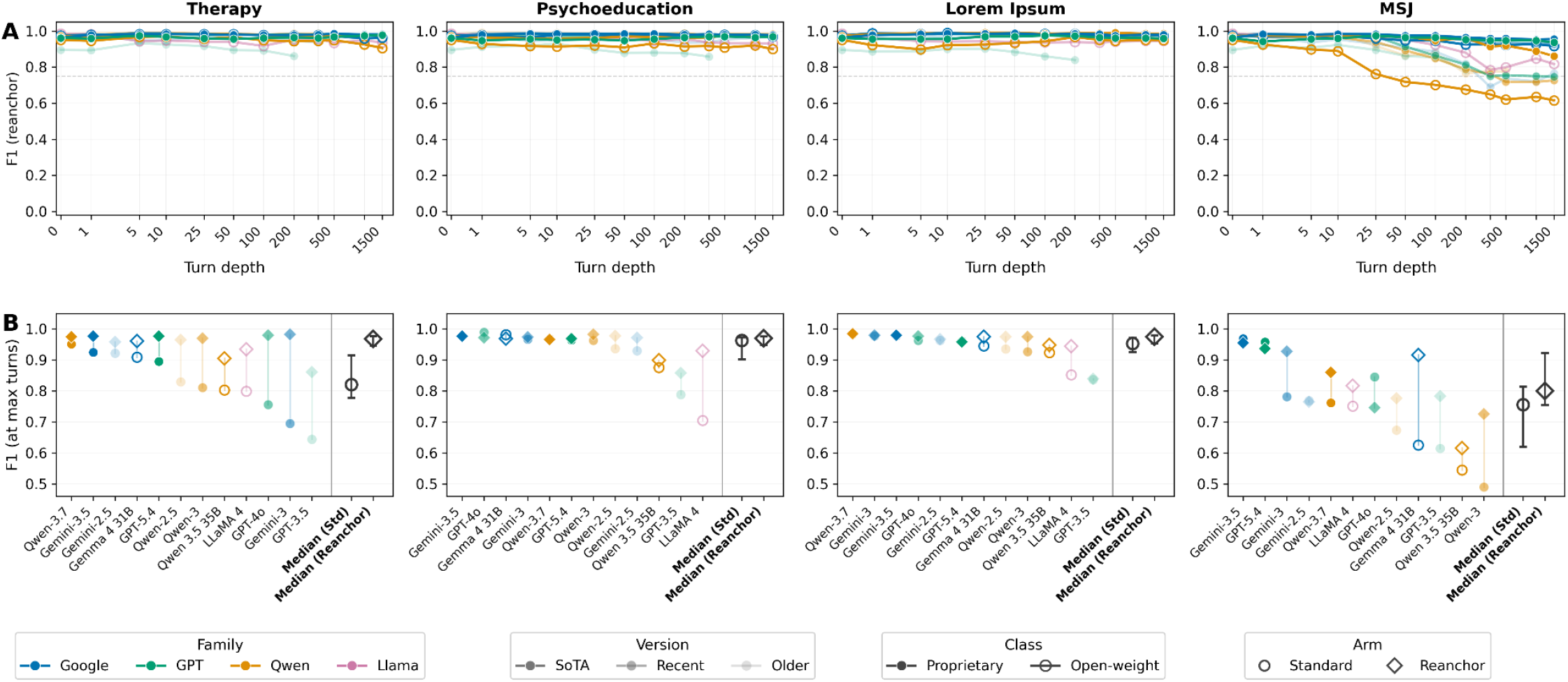
Effect of re-anchoring on top-performing models’ SI classification performance at maximum conversation depth. **A)** F1 score vs conversation turn depth (0–1500) with re-anchoring prompts for the same 12-model subset and conversation types as Figure 3. **B)** Paired F1 at each model’s maximum available turn depth, comparing performance without re-anchoring (circles) vs with re-anchoring (diamonds) at the same depth; models sorted within each panel by total F1 change. Maximum depth was 1500 turns for 11 of 12 models; GPT-3.5-turbo was context-window limited.

We next tested whether the content and direction of the preceding conversation, rather than its length alone, affects model classification. Conversation type explained variance beyond depth (likelihood-ratio χ^2^(3) = 187.1; p < 0.001): relative to therapy at matched depth, lorem ipsum (β = 0.034; p < 0.001) and psychoeducation (β = 0.017; p = 0.03) were positively associated with F1 score, while MSJ (β = −0.069; p < 0.001) was significantly negatively associated with F1 (**Table S8)**. MSJ degradation was strongly model-dependent (**Table S9**): F1 at maximum depth fell to 0.55 for Qwen 3.5 35B (from 0.93), 0.63 for Gemma 4 31B (from 0.98), and 0.77 for Gemini 2.5 Flash (from 0.96), whereas GPT-5.4 (0.96 to 0.96) and Gemini 3.5 Flash (0.96 to 0.97) were essentially unaffected.

### Re-anchoring

Restating the full classification instructions immediately before each potential SI statement raised F1 at each model’s maximum reached depth on therapy (median ΔF1, +0.12; p < 0.001), MSJ (+0.08; p = 0.04), and lorem ipsum (+0.009; p < 0.01) contexts, with a marginal effect on psychoeducation (+0.013; p = 0.052) (F**igure 4, Table S9**), and the benefit grew with conversation length (β = +0.006; p < 0.001) (**Table S10**). Median re-anchoring gain represented 89% of median standard-arm F1 loss from F1_0_ on therapy (median loss, 0.14) and 38% on MSJ (median loss, 0.23). In absolute terms, median F1 at maximum reached depth remained 0.01 below F1_0_ on therapy versus 0.14 on MSJ (**Table S9**). Depth-related loss accrued in therapeutic context was therefore largely recoverable by task reinstatement, whereas loss accrued under adversarial context was generally not. Results and protocol deviations all originally preregistered hypotheses are summarized in **Table S11**.

## Discussion

In this preregistered study assessing LLM and clinician detection of suicidal ideation in real psychotherapy transcripts, every LLM family tested showed conversation length and content-dependent degradation, whereas licensed clinicians showed no comparable decline. Notably, the best LLMs outperformed most clinicians in absolute terms, with eight of nine proprietary models exceeding clinician performance at 200 turns. These results provide direct experimental support for a distinction recently proposed in mental health AI safety between acute, single-exchange failures (type I harms) and insidious harms that accrue across extended interactions (type II harms)^8^. Models whose performance matches or exceeds clinicians on what is effectively a type I task nonetheless have that performance degrade over the conversational trajectories in which type II harms arise. To our knowledge, this is the largest multi-turn evaluation of safety-relevant LLM behavior conducted on real therapy transcripts, more than an order of magnitude beyond prior evaluations, which relied on simulated conversations of ~30 or fewer turns^16^. In addition, whereas many existing benchmarks score model outputs with other LLMs^22–25^, this study used licensed clinicians as the reference standard.

Consistent with prior work^26–30^, newer and larger models performed better. Parameter count and model version each predicted higher F1 scores within families, and proprietary models outperformed nearly all open-weight models, although the best open-weight models performed similarly to frontier models at early speaker turns.

Degradation was nonetheless detectable in every model family, indicating that length-dependent performance loss is a general property of current LLMs on this task rather than a limitation of smaller or older models.

Extending conversations to 1,500 speaker turns let us isolate conversational trajectory, ie the content and direction of the preceding conversation, from conversational length alone. Strikingly, trajectory, rather than length, drove observed performance loss. Lorem ipsum, semantically neutral filler conversation, and psychoeducation, direct instruction on appropriate suicidal-ideation assessment, produced at most modest losses, whereas many-shot jailbreaking (MSJ) adversarial conversation content produced steep declines. Although all top-performing models achieved excellent single-turn performance, their robustness to degradation varied widely. Two frontier models were essentially unaffected across 1,500 turns on all trajectories, while other models lost nearly half of their F1 under adversarial conditions. Critically, these behaviors are invisible to single- or few-turn evaluation benchmarks, which could overestimate performance at exactly the depths where users may be most vulnerable^10^.

Our re-anchoring experiments provide orthogonal support for these two distinct mechanisms of performance loss. For experiments using therapy, lorem ipsum, and psychoeducation conversations, re-presenting the classification instructions immediately prior to the potentially suicidal statement rescued performance, consistent with accumulating context displacing, rather than distorting, the model’s representation of the task. For the adversarial MSJ conversations, the same intervention failed to fully recover performance in most models: MSJ appears to warp the model’s understanding of the task rather than merely displacing the task instructions. This discrepancy is the signature of trajectory-dependent behavior: recoverability depended on what the conversation contained, not on its length.

These findings argue for restructuring safety evaluations of conversational AI in mental health around trajectories rather than individual user-model exchanges. Developers and evaluators should validate the specific model they deploy at realistic conversational depths on both therapeutic and adversarial trajectories, rather than inferring depth performance from few-turn benchmarks. Candidate mitigations should also be assessed with the same rigor and with their limits clearly stated. For example, in our work, restating safety-relevant instructions restored near-baseline performance for most therapeutic trajectories but not for adversarial ones. Limiting the total number of speaker turns reduced degradation in SI detection – making it attractive as a low-effort risk mitigation – but, without additional testing, may itself represent a new, independent hazard by interrupting continuity for precisely the users most engaged. Finally, deployed products embed models within routing, moderation, and memory systems that may attenuate or amplify these failure modes; it is the product in totality, not the model in isolation, whose safety ultimately matters.

This study has several limitations. First, caution and nuance are required when interpreting the clinician–LLM comparisons. This binary SI classification task is an artificial and deliberately narrow operationalization of one component of clinical safety evaluations. Actual clinical decisions are almost never dependent on whether a singular statement does or does not contain suicidal ideation. Instead, it’s the patient’s biopsychosocial gestalt – the longitudinal evolution and current state of their psychiatric and medical comorbidities and sociocultural background – combined with diagnostic interview, examination, and laboratory results that guides clinician judgment. Nor are clinicians commonly asked to evaluate a single statement for the presence of suicidal content at the end of a therapy transcript. That the best models outperformed most clinicians here is therefore not evidence that LLMs are ready to perform clinical risk assessment. The clinician arm also showed substantial inter-rater variability, underscoring that this detection task is difficult even for experienced clinicians. Second, the probe statements were synthetic. Although clinician-validated and stratified by severity and affective tone, they are not naturally occurring risk disclosures and may not reflect the semantic and syntactic language patterns, or their frequency distributions, from real patients. Third, the therapy transcripts were limited to five English-language psychotherapy transcripts with primarily depressive and anxious content from a single collection, and the extended depth experiments used concatenated transcripts rather than a single transcript from the same patient and session. Fourth, the clinician arm was small, and eight raters from a single institution may not reflect the broader range of clinical training and practice. Fifth, models were evaluated at temperature 0 without a sampling-parameter scan and therefore do not describe the response distribution introduced with varying model temperature. Finally, re-anchoring was tested only in the 12-model extension subset.

The central implication is that single or few-turn benchmarks can substantially mischaracterize the safety of conversational AI in mental health when the failure mode emerges from trajectory-dependent performance loss. Safety is not a fixed property of a model measured once, but a dynamic property of the model, the surrounding product, the user, their biopsychosocial state, and their interaction over time. Evaluations should therefore test sustained performance across realistic and adversarial trajectories before these systems are trusted in high-acuity settings where failure emerges gradually and where the users most at risk may be those who remain engaged the longest.

## Data Availability

The five raw psychotherapy transcripts cannot be redistributed under Alexander Street Press licensing terms and are therefore not included in the repository; to allow licensed users to regenerate the processed inputs, we provide the source document identifiers, the version-pinned processing script, and a SHA-256 manifest of the original files. Because the deposited analysis datasets are dialogue-free, the full statistical pipeline, tables, and figures can be reproduced from the repository without access to the licensed source transcripts deposited on OSF (DOI: doi.org/10.17605/osf.io/q8xam and doi.org/10.17605/osf.io/pvnbu) and GitHub (https://github.com/markkalinich/conversation-trajectory-multiturn-benchmark/).

https://www.doi.org/10.17605/osf.io/q8xam

https://www.doi.org/10.17605/osf.io/pvnbu

