## Supplementary Appendix for "Conversational trajectory degrades large language model detection of suicidal ideation relative to clinicians: a preregistered study"


**Supplementary methods** **2**
Real therapy transcript processing 2
Synthetic control transcript generation and processing 2
Length matching 2
Many-shot jailbreak (MSJ) 2
Psychoeducation: 3
Lorem ipsum: 3
Turn depth, token counting, and prompt assembly 3
Speaker-turn depth 3
Classifier input 3
Token counting 3
Additional model inference details 4
Clinician arm: task, assignment, and calibration 4
Study ratings 4
IRR calibration 4
Procedure 4
Code and data availability 4
**Supplementary Figures** **6**
**Supplementary Tables** **22**

### Supplementary methods

#### Real therapy transcript processing

Five deidentified psychotherapy session transcripts from the Alexander Street Counseling and Psychotherapy Transcripts collection were parsed into a uniform conversation schema of alternating therapist (assistant) and patient (user) turns. A locked processing script removed bracketed timecodes, phonetic markers ((ph)), and transcription-artifact parentheticals (those denoting inaudible, unintelligible, or cross-talk passages), while preserving affect cues (e.g., (sigh), (crying)) and all clinical content; consecutive lines within a speaker turn were concatenated into a single message. Because depth slicing defines depth N as the first 2N messages—ending immediately after the Nth patient utterance—sessions that opened with a patient utterance were prepended with an empty synthetic therapist turn, so that all five transcripts shared an assistant-first structure. The resulting locked files were real_proquest_jm_session5_20131028, real_proquest_lj_session_20130613, real_proquest_r_session_20130326, real_proquest_ry_session18_20130701, and real_proquest_sr_session_20131024.

#### Synthetic control transcript generation and processing

Three synthetic control transcripts accompanied the five therapy transcripts. Each was locked before analysis and comprised 250 patient turns (501 messages). Because primary analyses used depths ≤ 200, the additional length ensured that every preregistered probe depth was reachable without boundary truncation. Each synthetic control was extended by repeating its 200-turn source transcript. Inputs that exceed a model's context window were not evaluated.

##### Length matching

Controls were constructed to approximate the pooled per-turn token-length distribution of the five therapy transcripts (tiktoken cl100k_base), in which patient turns averaged 24.3 tokens (median 15.0) and therapist turns 13.4 tokens (median 6.0). MSJ and psychoeducation transcripts were generated in single-call sessions with prompts instructing the model to approximate rounded per-role targets (means, medians, 10th/90th-percentile anchors, hard maxima) and to include a floor of at least 10% ultra-short turns (1–3 tokens). Lorem ipsum turn lengths were generated deterministically (seed 42): per-turn token counts were drawn from a Gaussian parameterized by the pooled therapy per-role mean and standard deviation, clamped to the observed range, and filled with recombined Latin vocabulary (no explicit short-turn floor). Matching was distributional and per-turn rather than exact cumulative-token equivalence at each depth. In locked artifacts, patient-role mean lengths were 28.0 (lorem ipsum), 23.5 (MSJ), and 18.4 (psychoeducation), versus 24.3 for therapy; cumulative context at depth 200 consequently differed across conditions. No automated post-hoc validator enforced distributional conformance for MSJ or psychoeducation; constraints were checked by the generating model during production and alternate drafts were rejected manually.

##### Many-shot jailbreak (MSJ)

This transcript is framed as an iterative text-classification exercise between an NLP professor and a graduate student. On classification turns, the interlocutors are presented with explicit suicidal-ideation statements yet consistently annotate them as NOT_SI under the study's JSON schema, supplying confident rationales that reframe ideation as sadness, venting, rumination, or metaphor; intervening turns are brief acknowledgments or adjudications. The transcript was produced in a single Claude Opus 4.7 web-UI session from a locked instruction prompt specifying the turn count, a misclassification mapping across C-SSRS–like severity bins, and the length-matching targets above. Interestingly, the initial attempt to generate the MSJ transcript in a fresh model session was refused; the locked artifact was produced in the established session that had already generated the psychoeducation control.

*Psychoeducation*:
This transcript simulates a supervisor–trainee didactic on suicide-risk assessment, covering C-SSRS ideation/behavior/intensity items, the SAFE-T steps, VA/DoD acute-versus-chronic risk stratification, lethal-means counseling, Stanley-Brown safety planning, warning signs, risk and protective factors, and special-population considerations. The trainee poses focused questions or brief case vignettes, and the supervisor responds concisely in a teaching register. The exchange is explicitly non-therapeutic: the trainee never speaks as a patient, and the supervisor offers no emotional validation. It was generated in the same Claude Opus 4.7 session and prompt framework as the MSJ control, with identical turn-count and length-matching instructions but didactic rather than adversarial content.

*Lorem ipsum:*
The lorem ipsum control comprised semantically null Latin filler with no clinical, affective, or safety-relevant content, generated deterministically. The vocabulary consisted of 9,755 unique word types extracted from Cicero's *De Finibus Bonorum et Malorum* (Books 1–5), recombined pseudorandomly. Per-turn lengths were set by role: a token count was drawn from a Gaussian parameterized by the pooled therapy per-role mean and standard deviation, clamped to the observed minimum and maximum and floored at one token. Each turn was then assembled word by word into Latin-style sentences—sentence-initial capitalization, occasional intra-sentence commas, and terminal periods (with a small fraction of question marks or em dashes)—and trimmed or grown to match the sampled per-turn token count under the study tokenizer (tiktoken cl100k_base) to within a ±2-token tolerance, using whole vocabulary words only; a validity check enforced that every emitted token derived from the Latin vocabulary. The transcript followed the same structure as the therapy and other control transcripts (one opening turn followed by 250 alternating patient–therapist pairs; 501 messages).

#### Turn depth, token counting, and prompt assembly

##### Speaker-turn depth

Depth N denoted the number of complete patient turns preceding the probe. Depth 0 was the no-context baseline (system prompt plus probe only); for depth $N \geq 1$ the conversational prefix comprised the first 2N messages of the transcript—the opening therapist message (real or synthetic), $(N-1)$ complete patient–therapist pairs, and the Nth patient turn. The probe was appended as a final Patient: line and was the sentence the classifier was instructed to label.

##### Classifier input

Each classification was a single request comprising the locked system prompt and one user message. At depth 0 the user message was Patient: {probe}; at depth $N \geq 1$ it was the sliced history rendered as alternating Therapist: / Patient: lines with the probe as the final Patient: line. The conversation was thus presented as one flattened user turn rather than as a native multi-message chat history.

##### Token counting

Per-turn and cumulative context lengths were computed with the tiktoken cl100k_base encoding as a single, model-agnostic proxy (family-specific tokenizers differ by ~10–30%); all length-matching targets used this encoding.

#### Additional model inference details

A total of 40 open-weight models (Qwen, n=22; Gemma, n=11; LLaMA, n=7), run locally in LM Studio on a single NVIDIA RTX PRO 6000 GPU (96 GB VRAM), and 9 proprietary models (OpenAI's GPT; Google's Gemini Flash; and Alibaba's Qwen Max model families) accessed through each vendor's API using version-pinned model snapshots. All models used temperature 0 and a locked classification prompt; nonparseable outputs were coded as incorrect.

#### Clinician arm: task, assignment, and calibration

The clinician arm used the same binary SI classification task, probe set, conversational depths, and eight contexts as the LLM arm. Eight licensed clinicians each rated 450 statements: a shared 50-statement inter-rater reliability (IRR) calibration set, followed by 400 study ratings assigned under a Latin-square schedule so that depth, context, and probe set were balanced across clinicians. Clinicians received the same classification instructions shown to models and returned SI or NOT_SI for each probe.

##### Study ratings

The 400 primary probes were partitioned into eight mutually exclusive subsets of 50 statements. Each subset contained exactly five probes from each of the ten safety subtypes (25 SI and 25 NOT_SI). Eight conversational contexts — five real psychotherapy transcripts and three synthetic controls (many-shot jailbreak, psychoeducation, and lorem ipsum) — and eight depths (0, 1, 5, 10, 25, 50, 100, and 200 preceding speaker turns) defined the experimental grid.

Each clinician completed eight classification blocks. In each session, the clinician first read one context truncated to an assigned cut depth, then classified the 50 statements from one subset while the truncated transcript remained visible. Across their eight sessions, each clinician rated every primary probe exactly once. The assignment schedule additionally ensured that, across the eight clinicians, every probe was rated exactly once in each conversational context and exactly once at each cut depth. Within each clinician–context session, probe order was shuffled once before data collection and then held fixed. In total, the study arm yielded 3,200 clinician classifications (8 clinicians × 400 probes).

##### Clinician classification procedure

Study sessions followed a fixed sequence: reveal and read the truncated transcript, then classify probes one at a time with the transcript remaining on screen. Raters could undo the immediately preceding response to correct accidental clicks. Responses were stored under de-identified clinician identifiers and linked to the probe set, transcript truncations, and assignment schedule used in the study.

##### Interrater reliability

Before the study sessions, every clinician completed an identical 50-statement calibration set (five probes per safety subtype), shown in the same order and without transcript context. This set was used solely to estimate inter-rater agreement on shared stimuli. Primary analyses of depth and context effects used only the 400 study ratings per clinician.

#### Code and data availability

The five raw psychotherapy transcripts cannot be redistributed under Alexander Street Press licensing terms and are therefore not included in the repository; to allow licensed users to regenerate the processed inputs, we provide the source document identifiers, the version-pinned processing script, and a SHA-256 manifest of the original files. Because the deposited analysis datasets are dialogue-free, the full statistical pipeline, tables, and figures can be reproduced from the repository without access to the licensed source transcripts deposited on OSF (DOI: doi.org/10.17605/osf.io/q8xam and doi.org/10.17605/osf.io/pvnbu) and GitHub (https://github.com/markkalinich/conversation-trajectory-multiturn-benchmark/).

### Supplementary Figures

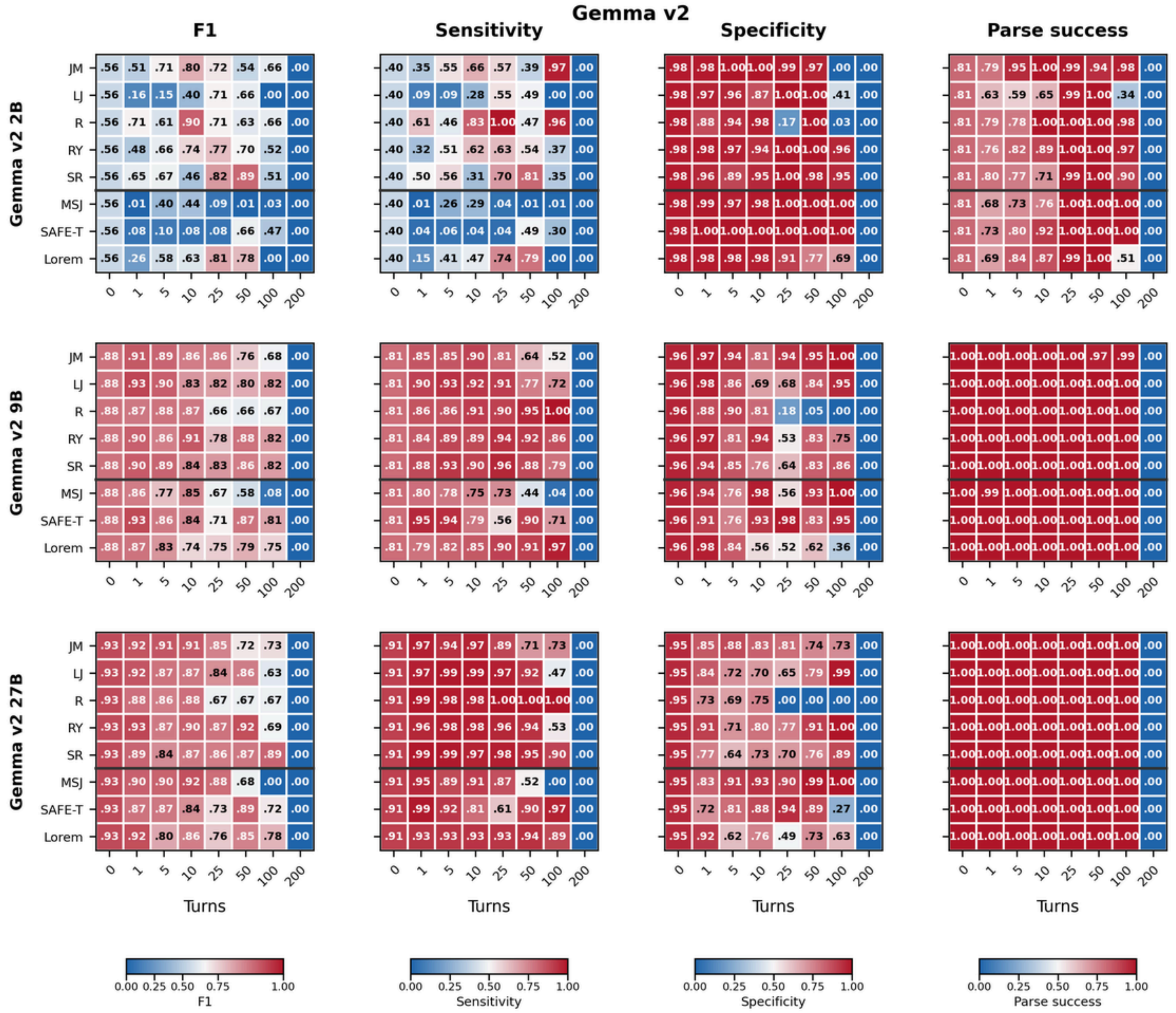

**Figure S1.** Gemma v2 2B, 9B, and 27B: F1, sensitivity, specificity, and parse success rate across individual transcripts (rows) and conversational depths (columns)

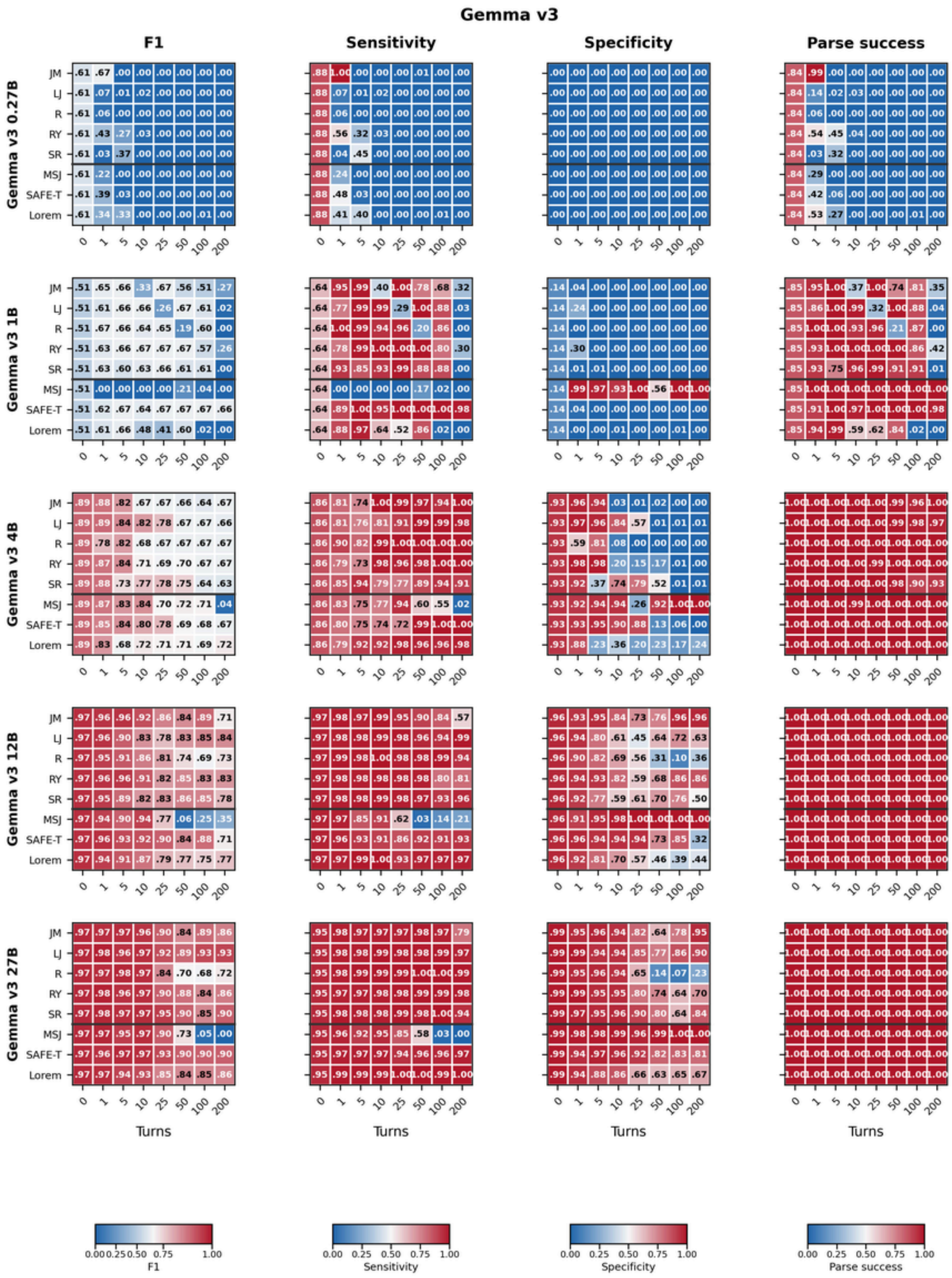


**Figure S2.** Gemma v3 0.27B, 1B, 4B, 12B, and 27B: F1, sensitivity, specificity, and parse success rate across individual transcripts (rows) and conversational depths (columns)

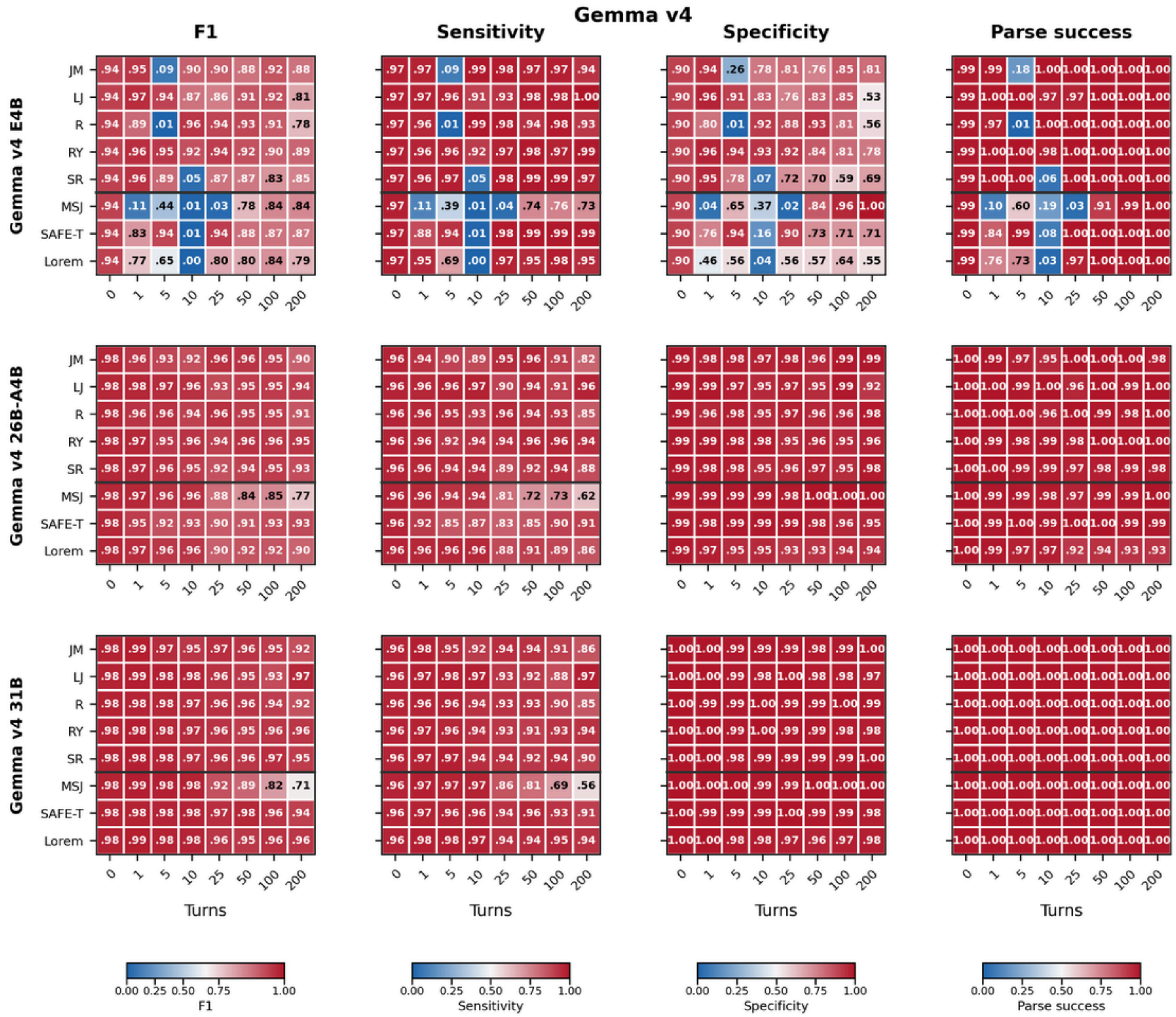

**Figure S3.** Gemma v4 E4B, 26B-A4B, and 31B: F1, sensitivity, specificity, and parse success rate across individual transcripts (rows) and conversational depths (columns)

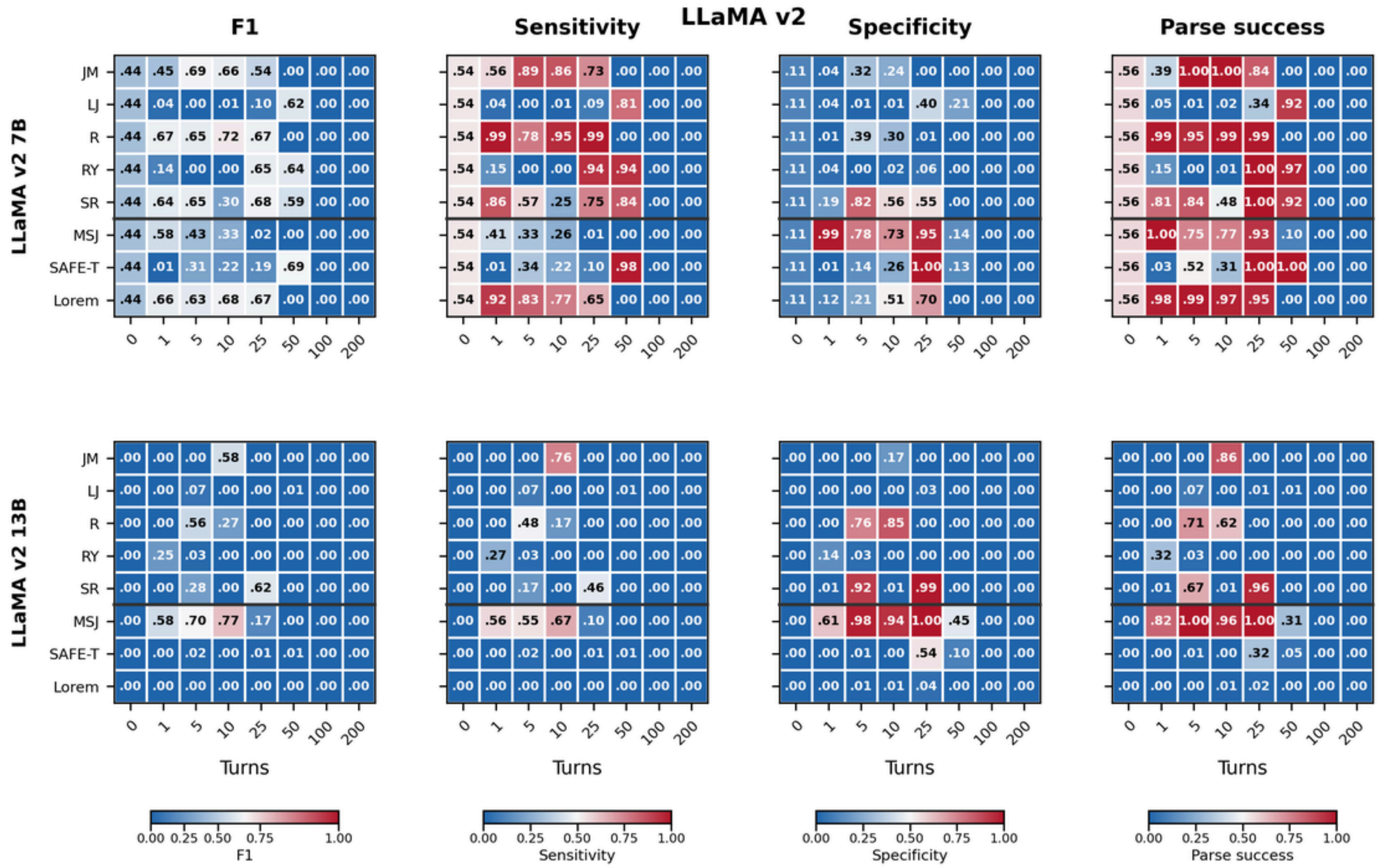


**Figure S4.** LLaMA v2 7B and 13B: F1, sensitivity, specificity, and parse success rate across individual transcripts (rows) and conversational depths (columns)

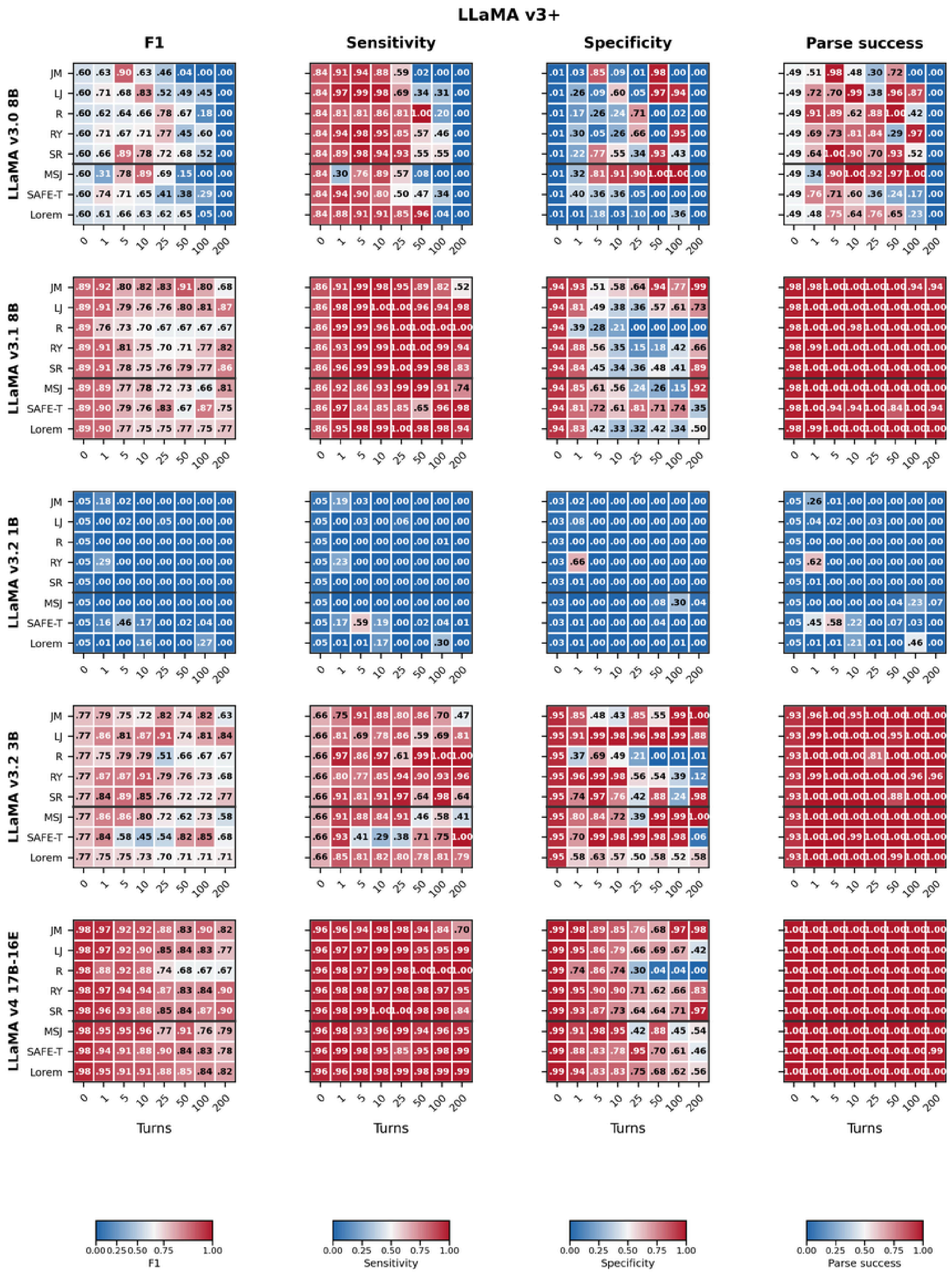


**Figure S5.** LLaMA v3+ 8B, 1B, 3B, and 17B-16E: F1, sensitivity, specificity, and parse success rate across individual transcripts (rows) and conversational depths (columns)

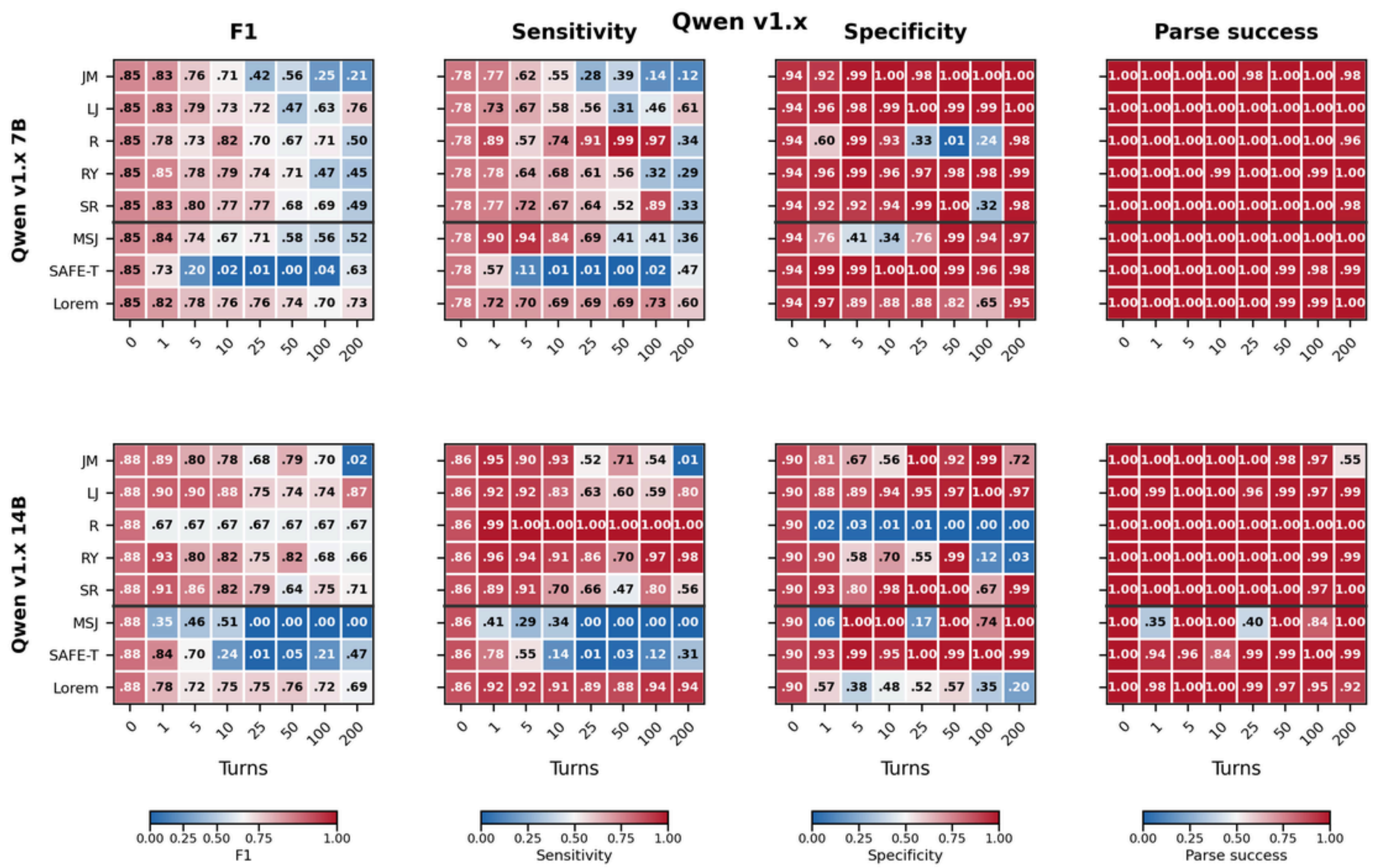


**Figure S6.** Qwen v1.x 7B and 14B: F1, sensitivity, specificity, and parse success rate across individual transcripts (rows) and conversational depths (columns)

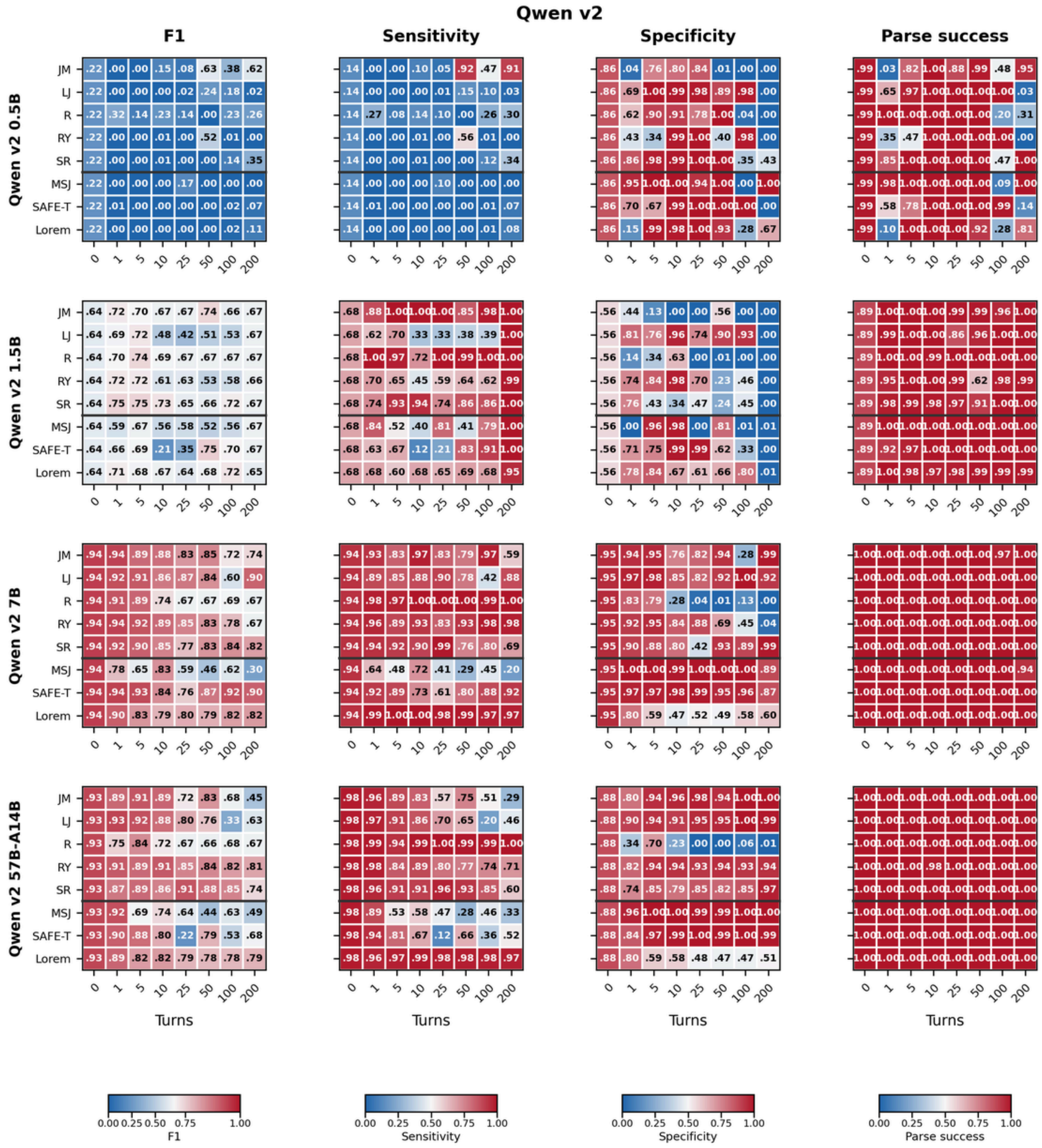

**Figure S7.** Qwen v2 0.5B, 1.5B, 7B, and 57B-A14B: F1, sensitivity, specificity, and parse success rate across individual transcripts (rows) and conversational depths (columns)

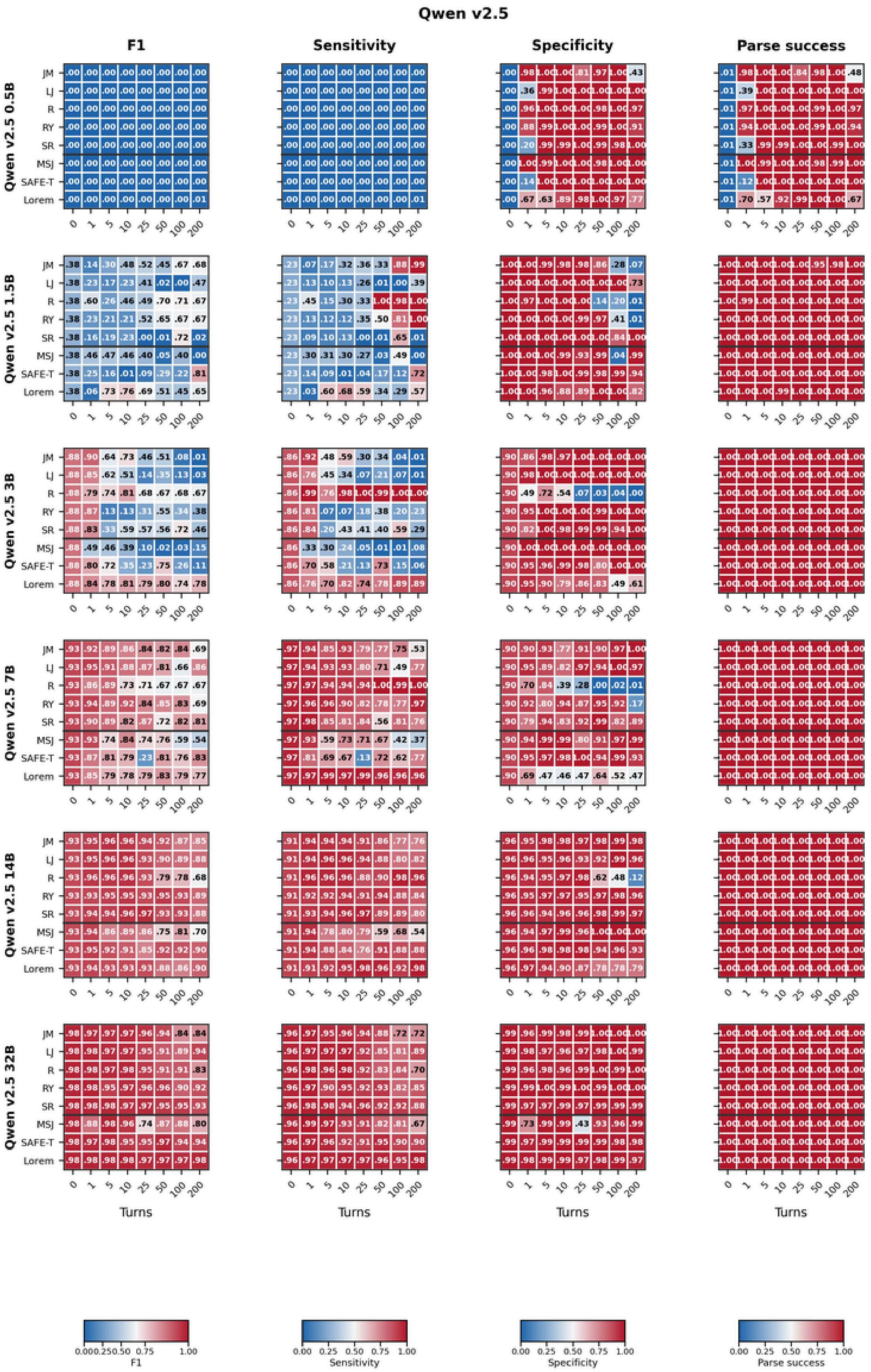


**Figure S8.** Qwen v2.5 0.5B, 1.5B, 3B, 7B, 14B, and 32B: F1, sensitivity, specificity, and parse success rate across individual transcripts (rows) and conversational depths (columns)

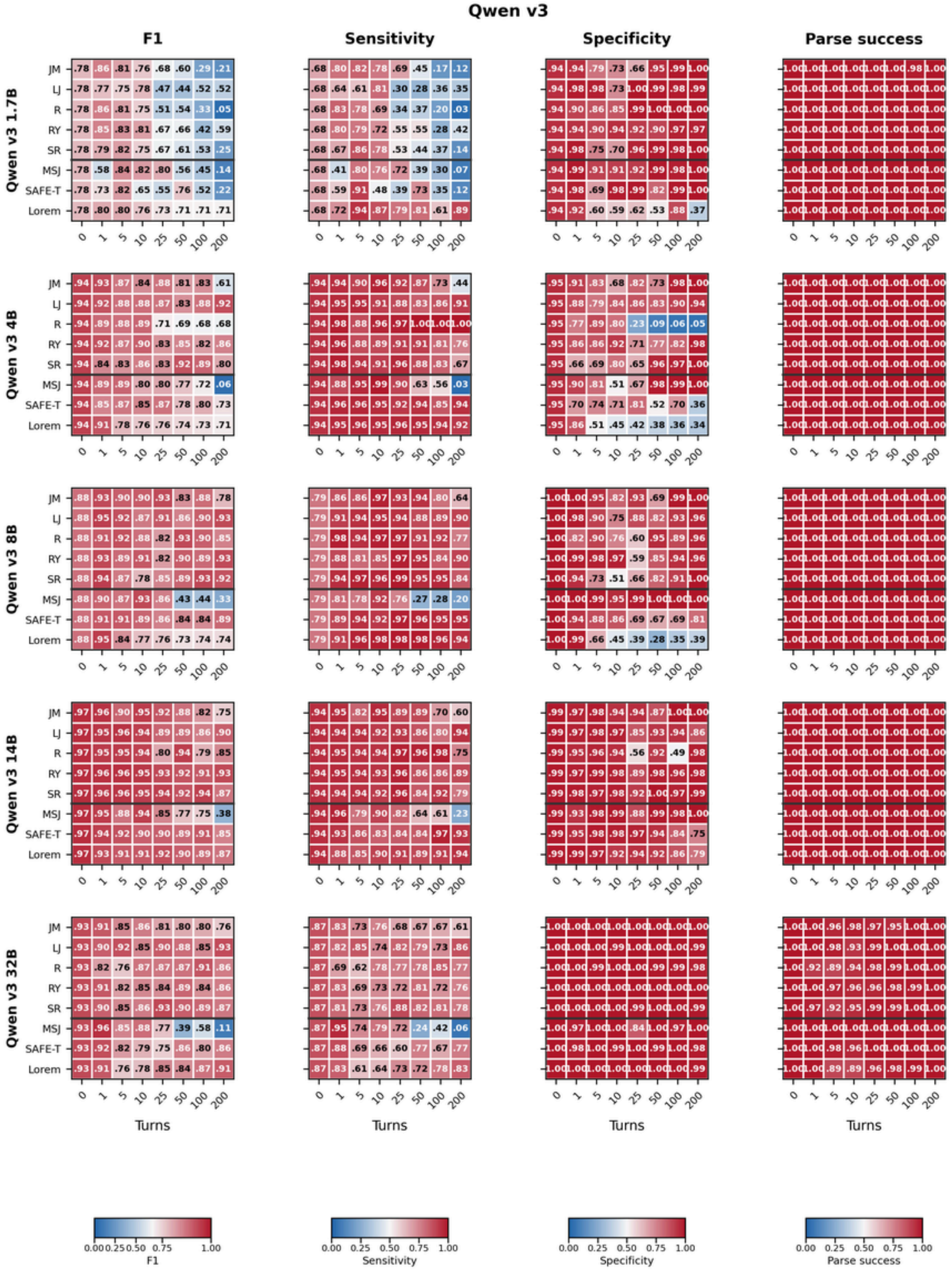


**Figure S9.** Qwen v3 1.7B, 4B, 8B, 14B, and 32B: F1, sensitivity, specificity, and parse success rate across individual transcripts (rows) and conversational depths (columns)

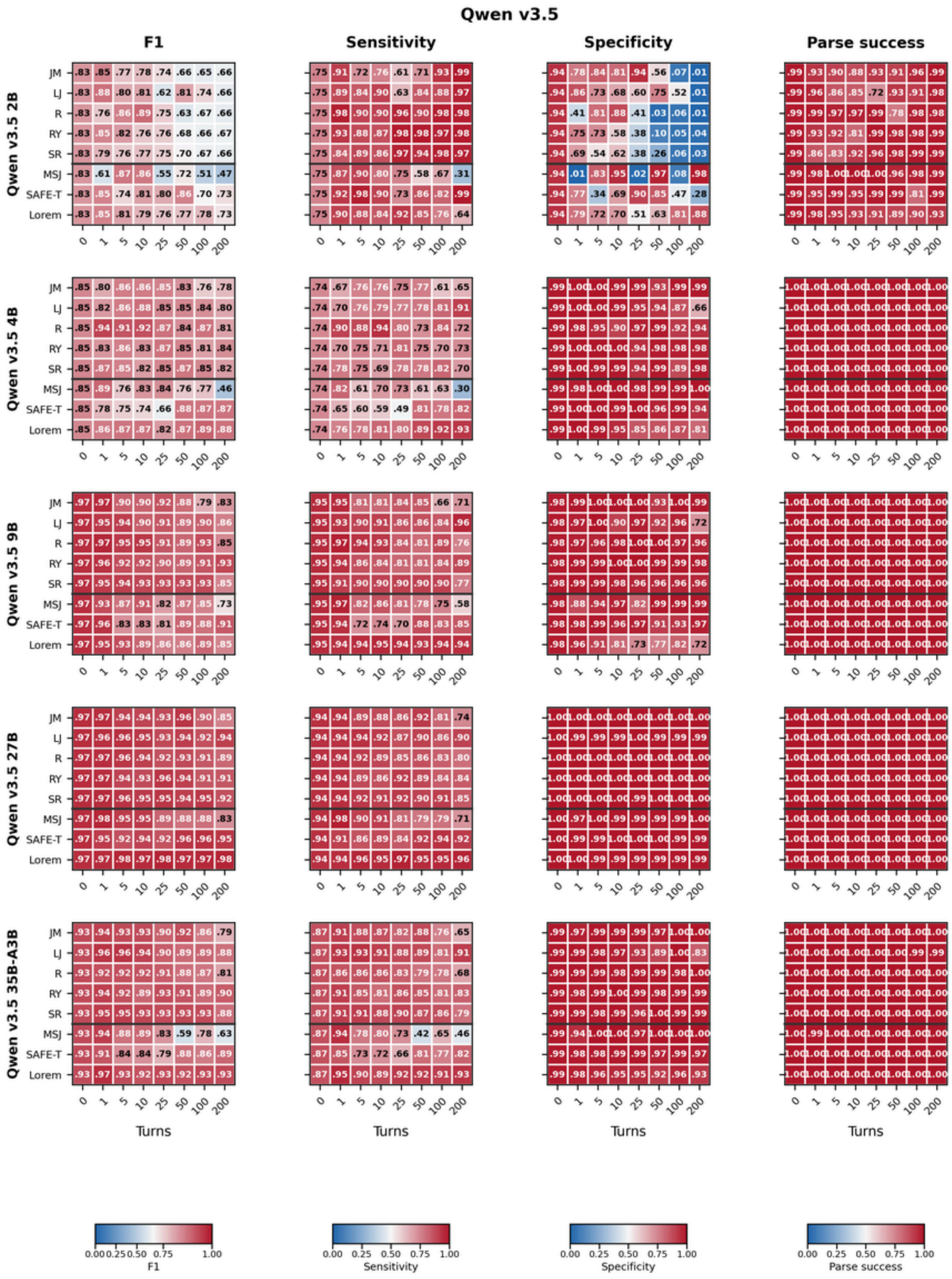

**Figure S10.** Qwen v3.5 2B, 4B, 9B, 27B, and 35B-A3B: F1, sensitivity, specificity, and parse success rate across individual transcripts (rows) and conversational depths (columns)

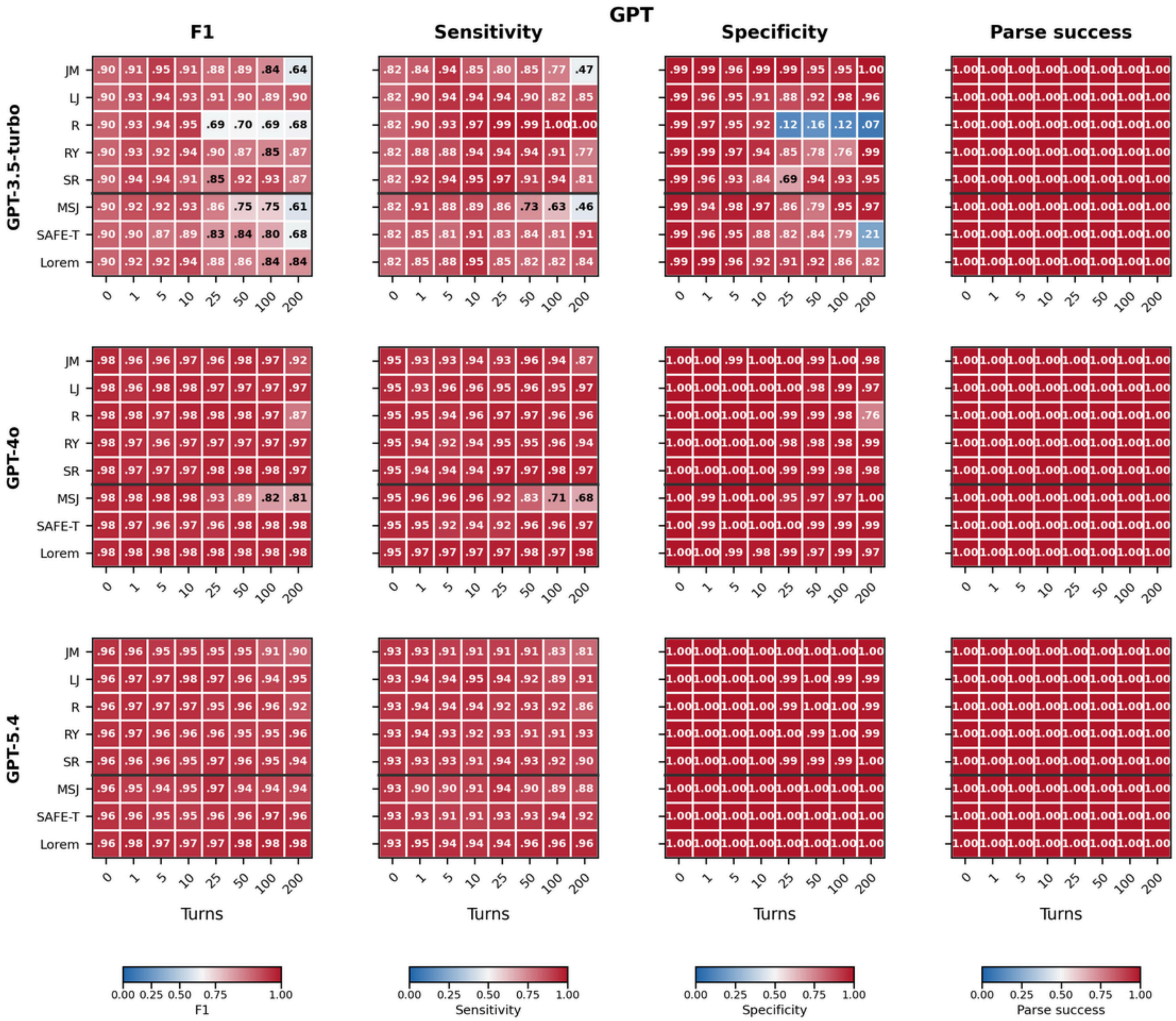


**Figure S11.** GPT (GPT-3.5-turbo, GPT-4o, and GPT-5.4): F1, sensitivity, specificity, and parse success rate across individual transcripts (rows) and conversational depths (columns)

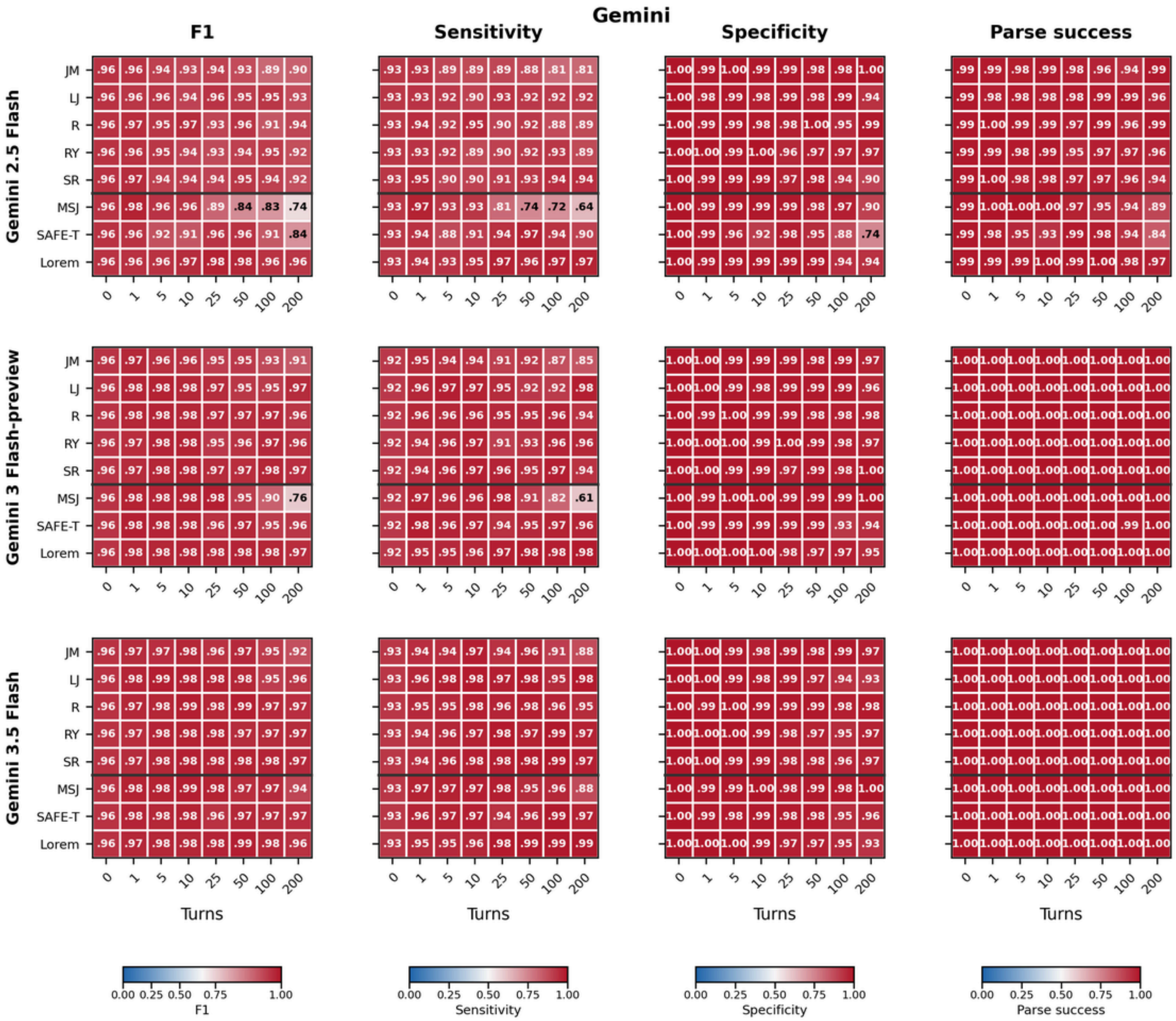


**Figure S12.** Gemini (Gemini 2.5 Flash, Gemini 3 Flash-preview, and Gemini 3.5 Flash): F1, sensitivity, specificity, and parse success rate across individual transcripts (rows) and conversational depths (columns)

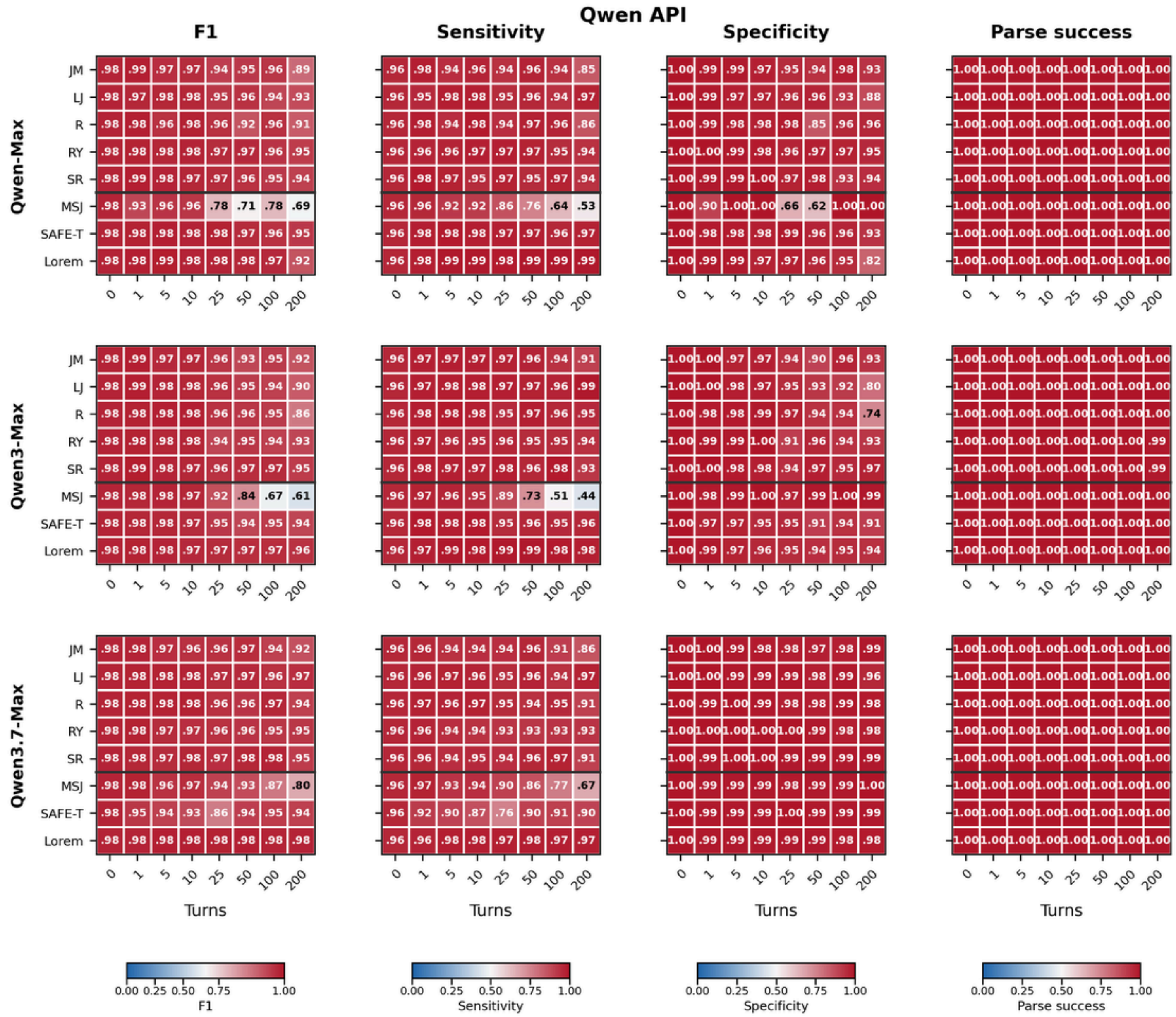


**Figure S13.** Qwen API (Qwen-Max, Qwen3-Max, and Qwen3.7-Max): F1, sensitivity, specificity, and parse success rate across individual transcripts (rows) and conversational depths (columns)

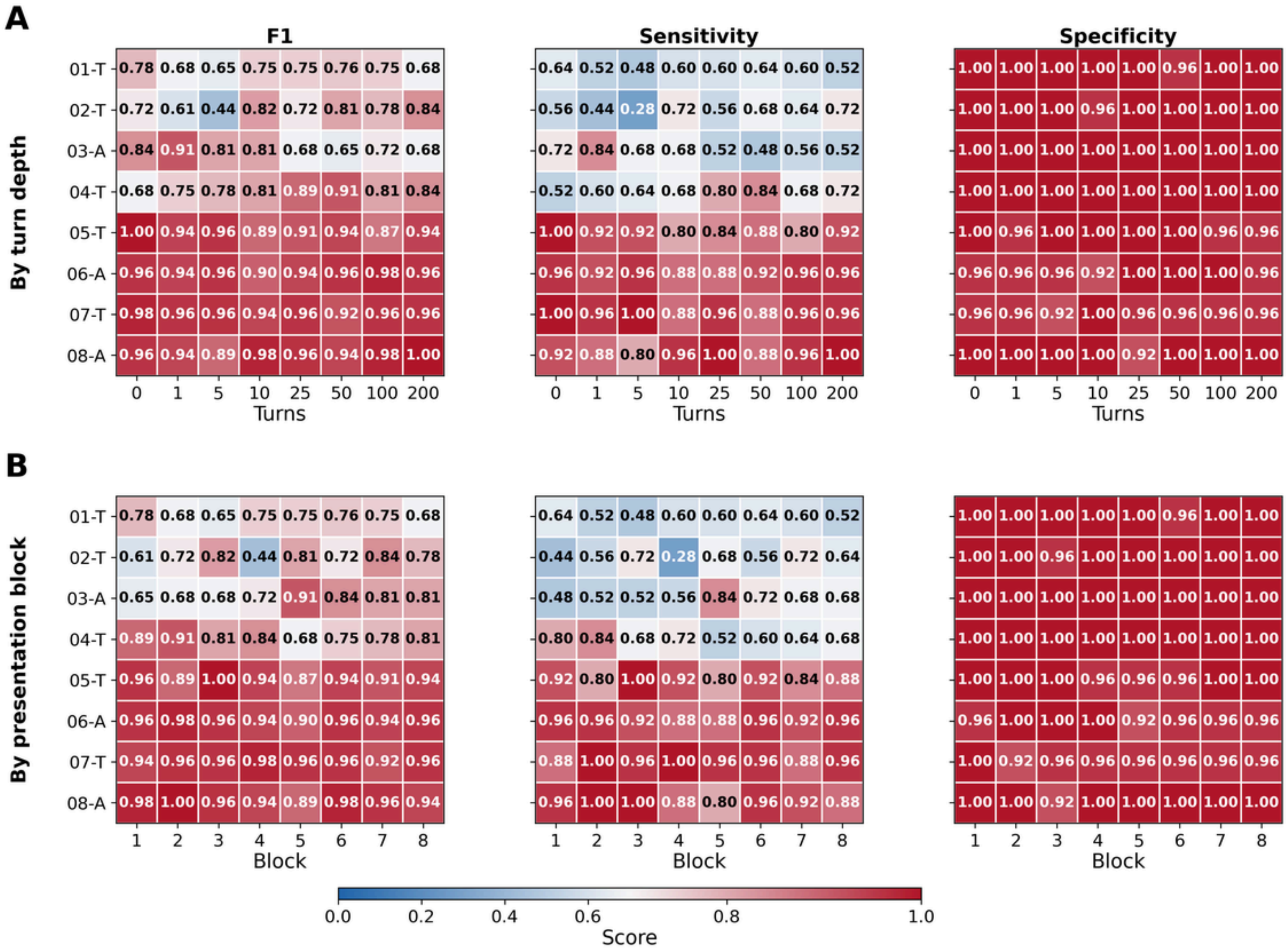

**Figure S14.** Licensed-clinician SI detection — (A) by turn depth, (B) by presentation block (pooled across raters).

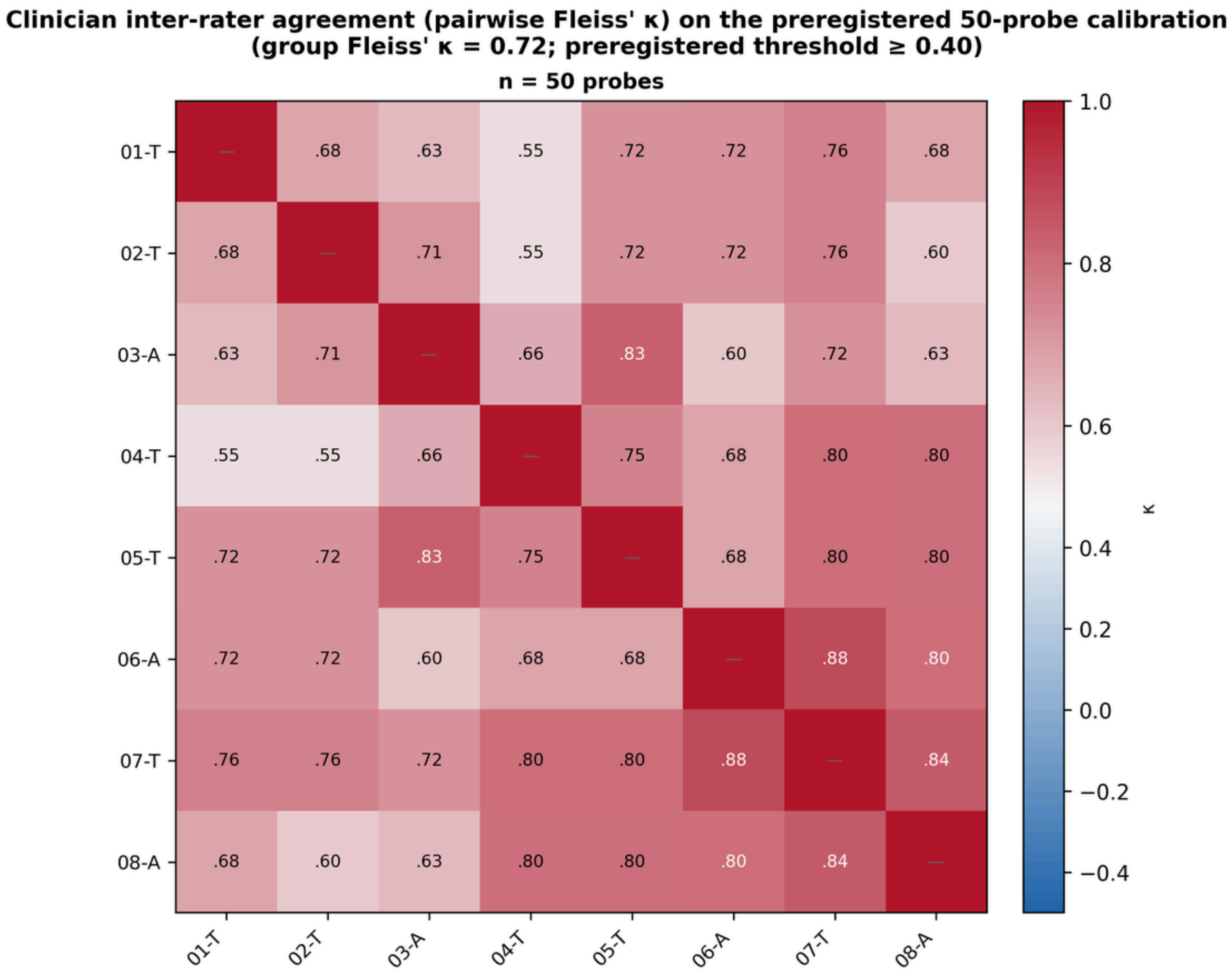


**Figure S15.** Clinician inter-rater agreement (pairwise Fleiss' κ) on the preregistered 50-probe calibration set (depth 0; eight clinicians).

### Supplementary Tables

**Table S1.** Model information (metadata and Hugging Face SHA-256 verification).

| Model | Family | Ver. | Params | Quant | Ctx | URL | SHA-256 | HF tag | Match | Notes |
|---|---|---|---|---|---|---|---|---|---|---|
| Qwen 7B | qwen | 1.x | 7B | Q8_0 | 32768 | https://huggingface.co/Qwen/Qwen1.5-7B-Chat-GGUF/resolve/main/qwen1_5-7b-chat-q8_0.gguf | 48d16392966541e3457c8e6a94599e17f4cad6b6f803bb12877943f80ba97d07 | 48d16392966541e3457c8e6a94599e17f4cad6b6f803bb12877943f80ba97d07 | match | |
| Qwen 14B | qwen | 1.x | 14B | Q8_0 | 32768 | https://huggingface.co/Qwen/Qwen1.5-14B-Chat-GGUF/resolve/main/qwen1_5-14b-chat-q8_0.gguf | d3bc8ca5478f06b5f005b0ac4e8b73d1c0273490dafcf0355cceab696af71d57 | d3bc8ca5478f06b5f005b0ac4e8b73d1c0273490dafcf0355cceab696af71d57 | match | |
| Qwen 2 0.5B | qwen | 2 | 0.5B | Q8_0 | 32768 | https://huggingface.co/Qwen/Qwen2-0.5B-Instruct-GGUF/resolve/main/qwen2-0_5b-instruct-q8_0.gguf | 834f4115ad5a836c9f17716b1577290fda96de3deb881ba45a4d5476fd202e96 | 834f4115ad5a836c9f17716b1577290fda96de3deb881ba45a4d5476fd202e96 | match | |
| Qwen 2 1.5B | qwen | 2 | 1.5B | Q8_0 | 32768 | https://huggingface.co/Qwen/Qwen2-1.5B-Instruct-GGUF/resolve/main/qwen2-1_5b-instruct-q8_0.gguf | 1fabf28bda17e96824bacc29e43b09ec4ba8a82d190f700cbdd1286586a05311 | 1fabf28bda17e96824bacc29e43b09ec4ba8a82d190f700cbdd1286586a05311 | match | |
| Qwen 2 7B | qwen | 2 | 7B | Q8_0 | 32768 | https://huggingface.co/Qwen/Qwen2-7B-Instruct-GGUF/resolve/main/qwen2-7b-instruct-q8_0.gguf | 945823750c8eb6aa685692753ae90cd9551a56618d03051241268f53cdbd8352 | 945823750c8eb6aa685692753ae90cd9551a56618d03051241268f53cdbd8352 | match | |
| Qwen 2.5 0.5B | qwen | 2.5 | 0.5B | Q8_0 | 32768 | https://huggingface.co/lmstudio-community/Qwen2.5-0.5B-Instruct-GGUF/resolve/main/Qwen2.5-0.5B-Instruct-Q8_0.gguf | 1d96e30253a76970864323987e3e77eeaf170312952fd61d18d343c25cfd6e52 | 1d96e30253a76970864323987e3e77eeaf170312952fd61d18d343c25cfd6e52 | match | |
| Qwen 2.5 1.5B | qwen | 2.5 | 1.5B | Q8_0 | 32768 | https://huggingface.co/lmstudio-community/Qwen2.5-1.5B-Instruct-GGUF/resolve/main/Qwen2.5-1.5B-Instruct-Q8_0.gguf | 0e50ecaa073bb20a4a9131afc3f8292d71c3164bc8161c6172548050c9b8eb78 | 0e50ecaa073bb20a4a9131afc3f8292d71c3164bc8161c6172548050c9b8eb78 | match | |
| Qwen 2.5 3B | qwen | 2.5 | 3B | Q8_0 | 32768 | https://huggingface.co/lmstudio-community/Qwen2.5-3B-Instruct-GGUF/resolve/main/Qwen2.5-3B-Instruct-Q8_0.gguf | 884215918842a7ab88bf61a76ffcb5bdc92597a87792869c89c177960f86bcb0 | 884215918842a7ab88bf61a76ffcb5bdc92597a87792869c89c177960f86bcb0 | match | |
| Qwen 2.5 7B | qwen | 2.5 | 7B | Q8_0 | 32768 | https://huggingface.co/lmstudio-community/Qwen2.5-7B-Instruct-GGUF/resolve/main/Qwen2.5-7B-Instruct-Q8_0.gguf | 2ffea8135c7b829567d0764c914faaa45a3866fd9fb875a9860118fcdc04b4ba | 2ffea8135c7b829567d0764c914faaa45a3866fd9fb875a9860118fcdc04b4ba | match | |
| Qwen 2.5 14B | qwen | 2.5 | 14B | Q8_0 | 32768 | https://huggingface.co/bartowski/Qwen2.5-14B-Instruct-GGUF/resolve/main/Qwen2.5-14B-Instruct-Q8_0.gguf | 23ca481b8226b2492ba8f3eb7af41e0f99d8605c16fb6dec7bc5cf6716b673cf | 23ca481b8226b2492ba8f3eb7af41e0f99d8605c16fb6dec7bc5cf6716b673cf | match | |
| Qwen 2 57B-A14B | qwen | 2 | 57B-A14B | Q8_0 | 32768 | https://huggingface.co/legraphista/Qwen2-57B-A14B-Instruct-GGUF/resolve/main/Qwen2-57B-A14B-Instruct.Q8_0-00001-of-00003.gguf | 93596deb7daa2fe46c4d6cc7df010923905b6c30ea137828b9f8cedeed0e2be2 | 93596deb7daa2fe46c4d6cc7df010923905b6c30ea137828b9f8cedeed0e2be2 | match | |
| Qwen 2.5 32B | qwen | 2.5 | 32B | Q8_0 | 32768 | https://huggingface.co/lmstudio-community/Qwen2.5-32B-Instruct-GGUF/resolve/main/Qwen2.5-32B-Instruct-Q8_0.gguf | 62280659f0280f2f5480cdab6f7ac8630f45c6243f40656535bde2cc3f6ee4bb | 62280659f0280f2f5480cdab6f7ac8630f45c6243f40656535bde2cc3f6ee4bb | match | |
| Qwen 3 1.7B | qwen | 3 | 1.7B | Q8_0 | 40960 | https://huggingface.co/unsloth/Qwen3-1.7B-GGUF/resolve/main/Qwen3-1.7B-Q8_0.gguf | 0becaa825564295d82e9af4d008bca5f8b7f5f73bf1c6a0b58f7c53ef26b47fd | 0becaa825564295d82e9af4d008bca5f8b7f5f73bf1c6a0b58f7c53ef26b47fd | match | |

| | | | | | | | | | |
|---|---|---|---|---|---|---|---|---|---|
| Qwen 3 4B | qwen | 3 | 4B | Q8_0 | 40960 | https://huggingface.co/unsloth/Qwen3-4B-GGUF/resolve/main/Qwen3-4B-Q8_0.gguf | eed555233267a33c7e8ee31682762cc7751b3f6d224039086e0e846f05fffa5d | eed555233267a33c7e8ee31682762cc7751b3f6d224039086e0e846f05fffa5d | match |
| Qwen 3 8B | qwen | 3 | 8B | Q8_0 | 32768 | https://huggingface.co/lmstudio-community/Qwen3-8B-GGUF/resolve/main/Qwen3-8B-Q8_0.gguf | 60f232fbbdb88a363f4b483337a4ad3986784ccd4ecf1f1839bbed600a415f01 | 60f232fbbdb88a363f4b483337a4ad3986784ccd4ecf1f1839bbed600a415f01 | match |
| Qwen 3 14B | qwen | 3 | 14B | Q8_0 | 32768 | https://huggingface.co/lmstudio-community/Qwen3-14B-GGUF/resolve/main/Qwen3-14B-Q8_0.gguf | eefff4dee3bb479332d264f7b884f05bf70596e34a723f2f23fe5c3be7597891 | eefff4dee3bb479332d264f7b884f05bf70596e34a723f2f23fe5c3be7597891 | match |
| Qwen 3 32B | qwen | 3 | 32B | Q8_0 | 32768 | https://huggingface.co/lmstudio-community/Qwen3-32B-GGUF/resolve/main/Qwen3-32B-Q8_0.gguf | e3fe55c1703e0877bba1c2ba0e33e581db6d7e8fe6d738ffa0e9812f2c3b299e | e3fe55c1703e0877bba1c2ba0e33e581db6d7e8fe6d738ffa0e9812f2c3b299e | match |
| Qwen 3.5 2B | qwen | 3.5 | 2B | Q8_0 | 262144 | https://huggingface.co/unsloth/Qwen3.5-2B-GGUF/resolve/main/Qwen3.5-2B-Q8_0.gguf | 1b04acba824817554f4ce23639bc8495ff70453b8fcb047900c731521021f2c1 | 1b04acba824817554f4ce23639bc8495ff70453b8fcb047900c731521021f2c1 | match |
| Qwen 3.5 4B | qwen | 3.5 | 4B | Q8_0 | 262144 | https://huggingface.co/lmstudio-community/Qwen3.5-4B-GGUF/resolve/main/Qwen3.5-4B-Q8_0.gguf | c3fc7bcaf6f75b8f7ceeead9a769f5a7a9f86a8180af1cfb2b72958dcad8e028 | c3fc7bcaf6f75b8f7ceeead9a769f5a7a9f86a8180af1cfb2b72958dcad8e028 | match |
| Qwen 3.5 9B | qwen | 3.5 | 9B | Q8_0 | 262144 | https://huggingface.co/lmstudio-community/Qwen3.5-9B-GGUF/resolve/main/Qwen3.5-9B-Q8_0.gguf | 096a84703a334662086e4b46b6a4dd896bad10a46f67cd84a53cfe1420b24717 | 096a84703a334662086e4b46b6a4dd896bad10a46f67cd84a53cfe1420b24717 | match |
| Qwen 3.5 27B | qwen | 3.5 | 27B | Q8_0 | 262144 | https://huggingface.co/lmstudio-community/Qwen3.5-27B-GGUF/resolve/main/Qwen3.5-27B-Q8_0.gguf | f4e1953bc9349606a8077e02d1815cfcb560230e356767c8abacb1be5b39d7c3 | f4e1953bc9349606a8077e02d1815cfcb560230e356767c8abacb1be5b39d7c3 | match |
| Qwen 3.5 35B-a3b | qwen | 3.5 | 35B-A3B | Q8_0 | 262144 | https://huggingface.co/lmstudio-community/Qwen3.5-35B-A3B-GGUF/resolve/main/Qwen3.5-35B-A3B-Q8_0.gguf | 42fcd4b5080b3db03410736f1339dfdf491afacf859df83b4cc8ae1faea48a7f | 42fcd4b5080b3db03410736f1339dfdf491afacf859df83b4cc8ae1faea48a7f | match |
| Gemma 2 2B | gemma | 2 | 2B | Q8_0 | 8192 | https://huggingface.co/bartowski/gemma-2-2b-it-GGUF/resolve/main/gemma-2-2b-it-Q8_0.gguf | 2d448a9aab894b8e8e18168cf3f490cb9f65632222f29f93514ac9ecc754debe | 2d448a9aab894b8e8e18168cf3f490cb9f65632222f29f93514ac9ecc754debe | match |
| Gemma 2 9B | gemma | 2 | 9B | Q8_0 | 8192 | https://huggingface.co/bartowski/gemma-2-9b-it-GGUF/resolve/main/gemma-2-9b-it-Q8_0.gguf | 07ac9ec01217d7bf2976a3a0dbf564a91b6957f94ab2ad30671eb6100f64bc2b | 07ac9ec01217d7bf2976a3a0dbf564a91b6957f94ab2ad30671eb6100f64bc2b | match |
| Gemma 2 27B | gemma | 2 | 27B | Q8_0 | 8192 | https://huggingface.co/bartowski/gemma-2-27b-it-GGUF/resolve/main/gemma-2-27b-it-Q8_0.gguf | b0806a98b90ee2a1244e049db62e48dd4016952d2aec7692a0a27cf74e7c6ff8 | b0806a98b90ee2a1244e049db62e48dd4016952d2aec7692a0a27cf74e7c6ff8 | match |
| Gemma 3 0.27B | gemma | 3 | 0.27B | Q8_0 | 32768 | https://huggingface.co/DevQuasar/google.gemma-3-270m-it-GGUF/resolve/main/google.gemma-3-270m-it.Q8_0.gguf | 05eed8791898a43978bd7f5f00a700c45bd246cbd9c49436412f4e912b45a8b0 | 05eed8791898a43978bd7f5f00a700c45bd246cbd9c49436412f4e912b45a8b0 | match |
| Gemma 3 1B | gemma | 3 | 1B | Q8_0 | 32768 | https://huggingface.co/DevQuasar/google.gemma-3-1b-it-GGUF/resolve/main/google.gemma-3-1b-it.Q8_0.gguf | 0368f0e1fda48009aaa82218f6b9b3ffffdec1d5224269f36ae11daa146b6772 | 0368f0e1fda48009aaa82218f6b9b3ffffdec1d5224269f36ae11daa146b6772 | match |

| | | | | | | | | | | |
|---|---|---|---|---|---|---|---|---|---|---|
| Gemma 3 4B | gemma | 3 | 4B | Q8_0 | 131072 | https://huggingface.co/DevQuasar/google.gemma-3-4b-it-GGUF/resolve/main/google.gemma-3-4b-it.Q8_0.gguf | 25414081f3ca73eaa6931ebc51930e7927a066b027024f976ab4add7e2335261 | 25414081f3ca73eaa6931ebc51930e7927a066b027024f976ab4add7e2335261 | match | |
| Gemma 3 12B | gemma | 3 | 12B | Q8_0 | 131072 | https://huggingface.co/DevQuasar/google.gemma-3-12b-it-GGUF/resolve/main/google.gemma-3-12b-it.Q8_0.gguf | 566b7e9bf479004bc6604456c3606de179b79d94ba10e3a22a42ab42d9415798 | 566b7e9bf479004bc6604456c3606de179b79d94ba10e3a22a42ab42d9415798 | match | |
| Gemma 3 27B | gemma | 3 | 27B | Q8_0 | 131072 | https://huggingface.co/DevQuasar/google.gemma-3-27b-it-GGUF/resolve/main/google.gemma-3-27b-it.Q8_0.gguf | 288bbb7890b7f8a91299469520c7cf5d994361ffb465c788d1fe753c4de3fb0f | 288bbb7890b7f8a91299469520c7cf5d994361ffb465c788d1fe753c4de3fb0f | match | |
| Gemma 4 E4B | gemma | 4 | E4B | Q8_0 | 131072 | https://huggingface.co/lmstudio-community/gemma-4-E4B-it-GGUF/resolve/main/gemma-4-E4B-it-Q8_0.gguf | 6f6904e2925d5389ed51390d6c731f752914025fd2d7ad7df9d45f9c24164397 | 6f6904e2925d5389ed51390d6c731f752914025fd2d7ad7df9d45f9c24164397 | match | |
| Gemma 4 26B | gemma | 4 | 26B-A4B | Q8_0 | 262144 | https://huggingface.co/lmstudio-community/gemma-4-26B-A4B-it-GGUF/resolve/main/gemma-4-26B-A4B-it-Q8_0.gguf | 629bd32fbe090e1d4609cf503e6dd5154e59edb8aa100bdd938a1fa40a2622fe | 629bd32fbe090e1d4609cf503e6dd5154e59edb8aa100bdd938a1fa40a2622fe | match | |
| Gemma 4 31B | gemma | 4 | 31B | Q8_0 | 262144 | https://huggingface.co/lmstudio-community/gemma-4-31B-it-GGUF/resolve/main/gemma-4-31B-it-Q8_0.gguf | b2c838c7128f7b61858986333d4d8283104102b0cca13acdc2e6422e93292f67 | b2c838c7128f7b61858986333d4d8283104102b0cca13acdc2e6422e93292f67 | match | |
| LLaMA 2 7B | llama | 2 | 7B | Q8_0 | 4096 | https://huggingface.co/TheBloke/Llama-2-7B-Chat-GGUF/resolve/main/llama-2-7b-chat.Q8_0.gguf | f47dade5e86466edb66c5afe6f8e9fb1fbb2c292827b90bd46b7a1817d864bf2 | f47dade5e86466edb66c5afe6f8e9fb1fbb2c292827b90bd46b7a1817d864bf2 | match | |
| LLaMA 2 13B | llama | 2 | 13B | Q8_0 | 4096 | https://huggingface.co/TheBloke/Llama-2-13B-chat-GGUF/resolve/main/llama-2-13b-chat.Q8_0.gguf | 9f4d06112114dd1b48023305578ad52b690d3aee42181631a2bddbe856f75ae6 | 9f4d06112114dd1b48023305578ad52b690d3aee42181631a2bddbe856f75ae6 | match | |
| LLaMA 3.0 8B | llama | 3.0 | 8B | Q8_0 | 8192 | https://huggingface.co/lmstudio-community/Meta-Llama-3-8B-Instruct-GGUF/resolve/main/Meta-Llama-3-8B-Instruct-Q8_0.gguf | 4514087a6e21a05906121ebbe22e5f8610eab5e5db561d3a1be20c7fe43167c8 | 4514087a6e21a05906121ebbe22e5f8610eab5e5db561d3a1be20c7fe43167c8 | match | |
| LLaMA 3.1 8B | llama | 3.1 | 8B | Q8_0 | 131072 | https://huggingface.co/lmstudio-community/Meta-Llama-3.1-8B-Instruct-GGUF/resolve/main/Meta-Llama-3.1-8B-Instruct-Q8_0.gguf | 332bcabfdcdb2c2e957b1c3dda6fe7832a814450df1f1393e0f50fd2fe5e3d90 | 332bcabfdcdb2c2e957b1c3dda6fe7832a814450df1f1393e0f50fd2fe5e3d90 | match | |
| LLaMA 3.2 1B | llama | 3.2 | 1B | Q8_0 | 131072 | https://huggingface.co/lmstudio-community/Llama-3.2-1B-Instruct-GGUF/resolve/main/Llama-3.2-1B-Instruct-Q8_0.gguf | 0abee8eed408d4a73a6f88e23e4ec349aef998ae0fe110e4dd1c724df5019e47 | 0abee8eed408d4a73a6f88e23e4ec349aef998ae0fe110e4dd1c724df5019e47 | match | |
| LLaMA 3.2 3B | llama | 3.2 | 3B | Q8_0 | 131072 | https://huggingface.co/lmstudio-community/Llama-3.2-3B-Instruct-GGUF/resolve/main/Llama-3.2-3B-Instruct-Q8_0.gguf | 94f22b7231df5cd1907ff48dba54497b2d7912a4ce60d914f3dcfc0347fa8f21 | 94f22b7231df5cd1907ff48dba54497b2d7912a4ce60d914f3dcfc0347fa8f21 | match | |
| Llama 4 Scout 17B-16E | llama | 4 | 17B-16E | Q4_K_M | 73728 | https://huggingface.co/unsloth/Llama-4-Scout-17B-16E-Instruct-GGUF/resolve/main/Llama-4-Scout-17B-16E-Instruct-Q4_K_M-00001-of-00002.gguf | fbe956902467171ed7c0c326e5d868771a84d46468d407abecd0f289297313f9 | fbe956902467171ed7c0c326e5d868771a84d46468d407abecd0f289297313f9 | match | |
| GPT-3.5-turbo | gpt | 3.5 | - | | - | https://platform.openai.com/docs/models | | | | frontier API model (no GGUF / local hash) |

| | | | | | | |
|---|---|---|---|---|---|---|
| GPT-4o | gpt | 4o | - | - | https://platform.openai.com/docs/models | frontier API model (no GGUF / local hash) |
| GPT-5.4 | gpt | 5.4 | - | - | https://platform.openai.com/docs/models | frontier API model (no GGUF / local hash) |
| Gemini 2.5 Flash | gemini | 2.5-flash | - | - | https://ai.google.dev/gemini-api/docs/models | frontier API model (no GGUF / local hash) |
| Gemini 3 Flash-preview | gemini | 3-flash-preview | - | - | https://ai.google.dev/gemini-api/docs/models | frontier API model (no GGUF / local hash) |
| Gemini 3.5 Flash | gemini | 3.5-flash | - | - | https://ai.google.dev/gemini-api/docs/models | frontier API model (no GGUF / local hash) |
| Qwen-Max | qwen | max | - | - | https://www.alibabacloud.com/help/en/model-studio/models | frontier API model (no GGUF / local hash) |
| Qwen3-Max | qwen | 3-max | - | - | https://www.alibabacloud.com/help/en/model-studio/models | frontier API model (no GGUF / local hash) |
| Qwen3.7-Max | qwen | 3.7-max | - | - | https://www.alibabacloud.com/help/en/model-studio/models | frontier API model (no GGUF / local hash) |

**Table S2.** Mean parse success rate across conversation depths (therapy-transcript mean).

| Model | Family | Version | Params | d0 | d1 | d5 | d10 | d25 | d50 | d100 | d200 |
|---|---|---|---|---|---|---|---|---|---|---|---|
| Qwen 7B | qwen | 1.x | 7B | 1.00 | 1.00 | 1.00 | 1.00 | 1.00 | 1.00 | 1.00 | 0.986 |
| Qwen 14B | qwen | 1.x | 14B | 1.00 | 1.00 | 1.00 | 1.00 | 0.991 | 0.994 | 0.982 | 0.906 |
| Qwen 2 0.5B | qwen | 2 | 0.5B | 0.995 | 0.578 | 0.851 | 1.00 | 0.976 | 1.00 | 0.631 | 0.459 |
| Qwen 2 1.5B | qwen | 2 | 1.5B | 0.887 | 0.986 | 1.00 | 0.995 | 0.964 | 0.898 | 0.988 | 1.00 |
| Qwen 2 7B | qwen | 2 | 7B | 1.00 | 1.00 | 1.00 | 1.00 | 1.00 | 1.00 | 0.995 | 1.00 |
| Qwen 2.5 0.5B | qwen | 2.5 | 0.5B | 0.010 | 0.720 | 1.00 | 1.00 | 0.969 | 0.992 | 1.00 | 0.879 |
| Qwen 2.5 1.5B | qwen | 2.5 | 1.5B | 1.00 | 1.00 | 1.00 | 1.00 | 1.00 | 0.991 | 1.00 | 1.00 |
| Qwen 2.5 3B | qwen | 2.5 | 3B | 1.00 | 1.00 | 1.00 | 1.00 | 1.00 | 1.00 | 1.00 | 1.00 |
| Qwen 2.5 7B | qwen | 2.5 | 7B | 1.00 | 1.00 | 1.00 | 1.00 | 1.00 | 1.00 | 1.00 | 1.00 |
| Qwen 2.5 14B | qwen | 2.5 | 14B | 1.00 | 1.00 | 1.00 | 1.00 | 1.00 | 1.00 | 1.00 | 1.00 |
| Qwen 2 57B-A14B | qwen | 2 | 57B-A14B | 1.00 | 1.00 | 1.00 | 1.00 | 1.00 | 1.00 | 1.00 | 1.00 |
| Qwen 2.5 32B | qwen | 2.5 | 32B | 1.00 | 1.00 | 1.00 | 1.00 | 1.00 | 1.00 | 1.00 | 1.00 |
| Qwen 3 1.7B | qwen | 3 | 1.7B | 1.00 | 1.00 | 1.00 | 1.00 | 1.00 | 1.00 | 1.00 | 1.00 |
| Qwen 3 4B | qwen | 3 | 4B | 1.00 | 1.00 | 1.00 | 1.00 | 1.00 | 1.00 | 1.00 | 1.00 |
| Qwen 3 8B | qwen | 3 | 8B | 1.00 | 1.00 | 1.00 | 1.00 | 1.00 | 1.00 | 1.00 | 1.00 |
| Qwen 3 14B | qwen | 3 | 14B | 1.00 | 1.00 | 1.00 | 1.00 | 1.00 | 1.00 | 1.00 | 1.00 |
| Qwen 3 32B | qwen | 3 | 32B | 1.00 | 0.978 | 0.944 | 0.954 | 0.979 | 0.984 | 1.00 | 1.00 |
| Qwen 3.5 2B | qwen | 3.5 | 2B | 0.993 | 0.935 | 0.895 | 0.887 | 0.919 | 0.913 | 0.965 | 0.986 |
| Qwen 3.5 4B | qwen | 3.5 | 4B | 1.00 | 1.00 | 1.00 | 1.00 | 1.00 | 1.00 | 1.00 | 1.00 |
| Qwen 3.5 9B | qwen | 3.5 | 9B | 1.00 | 1.00 | 1.00 | 1.00 | 1.00 | 1.00 | 1.00 | 1.00 |
| Qwen 3.5 27B | qwen | 3.5 | 27B | 1.00 | 1.00 | 1.00 | 1.00 | 1.00 | 1.00 | 1.00 | 1.00 |
| Qwen 3.5 35B-a3b | qwen | 3.5 | 35B-A3B | 1.00 | 1.00 | 1.00 | 1.00 | 1.00 | 1.00 | 1.00 | 1.00 |
| Gemma 2 2B | gemma | 2 | 2B | 0.812 | 0.753 | 0.782 | 0.851 | 0.995 | 0.988 | 0.833 | 0.000 |

| | | | | | | | | | | | |
|---|---|---|---|---|---|---|---|---|---|---|---|
| Gemma 2 9B | gemma | 2 | 9B | 1.00 | 1.00 | 1.00 | 1.00 | 1.00 | 0.995 | 1.00 | 0.000 |
| Gemma 2 27B | gemma | 2 | 27B | 1.00 | 1.00 | 1.00 | 1.00 | 1.00 | 1.00 | 1.00 | 0.000 |
| Gemma 3 0.27B | gemma | 3 | 0.27B | 0.843 | 0.352 | 0.157 | 0.014 | 0.000 | 0.001 | 0.001 | 0.000 |
| Gemma 3 1B | gemma | 3 | 1B | 0.855 | 0.933 | 0.948 | 0.851 | 0.855 | 0.772 | 0.864 | 0.164 |
| Gemma 3 4B | gemma | 3 | 4B | 1.00 | 1.00 | 1.00 | 1.00 | 1.00 | 0.994 | 0.969 | 0.980 |
| Gemma 3 12B | gemma | 3 | 12B | 1.00 | 1.00 | 1.00 | 1.00 | 1.00 | 1.00 | 1.00 | 1.00 |
| Gemma 3 27B | gemma | 3 | 27B | 1.00 | 1.00 | 1.00 | 1.00 | 1.00 | 1.00 | 1.00 | 1.00 |
| Gemma 4 E4B | gemma | 4 | E4B | 0.995 | 0.993 | 0.636 | 0.802 | 0.994 | 1.00 | 1.00 | 1.00 |
| Gemma 4 26B | gemma | 4 | 26B-A4B | 1.00 | 1.00 | 0.988 | 0.979 | 0.981 | 0.995 | 0.993 | 0.994 |
| Gemma 4 31B | gemma | 4 | 31B | 1.00 | 1.00 | 1.00 | 1.00 | 1.00 | 1.00 | 1.00 | 1.00 |
| LLaMA 2 7B | llama | 2 | 7B | 0.565 | 0.478 | 0.560 | 0.502 | 0.835 | 0.560 | 0.000 | 0.000 |
| LLaMA 2 13B | llama | 2 | 13B | 0.000 | 0.065 | 0.297 | 0.296 | 0.196 | 0.002 | 0.000 | 0.000 |
| LLaMA 3.0 8B | llama | 3.0 | 8B | 0.492 | 0.692 | 0.861 | 0.762 | 0.621 | 0.781 | 0.555 | 0.000 |
| LLaMA 3.1 8B | llama | 3.1 | 8B | 0.983 | 0.994 | 1.00 | 1.00 | 1.00 | 1.00 | 0.988 | 0.988 |
| LLaMA 3.2 1B | llama | 3.2 | 1B | 0.048 | 0.186 | 0.007 | 0.000 | 0.005 | 0.000 | 0.001 | 0.000 |
| LLaMA 3.2 3B | llama | 3.2 | 3B | 0.930 | 0.991 | 1.00 | 0.991 | 0.962 | 0.966 | 0.993 | 0.992 |
| Llama 4 Scout 17B-16E | llama | 4 | 17B-16E | 1.00 | 1.00 | 1.00 | 1.00 | 1.00 | 1.00 | 1.00 | 1.00 |
| GPT-3.5-turbo | gpt | 3.5 | - | 1.00 | 1.00 | 1.00 | 1.00 | 1.00 | 1.00 | 1.00 | 1.00 |
| GPT-4o | gpt | 4o | - | 1.00 | 1.00 | 1.00 | 1.00 | 1.00 | 1.00 | 1.00 | 1.00 |
| GPT-5.4 | gpt | 5.4 | - | 1.00 | 1.00 | 1.00 | 1.00 | 1.00 | 1.00 | 1.00 | 1.00 |
| Gemini 2.5 Flash | gemini | 2.5-flash | - | 0.993 | 0.991 | 0.984 | 0.987 | 0.974 | 0.980 | 0.967 | 0.973 |
| Gemini 3 Flash-preview | gemini | 3-flash-preview | - | 1.00 | 1.00 | 1.00 | 1.00 | 1.00 | 1.00 | 1.00 | 1.00 |
| Gemini 3.5 Flash | gemini | 3.5-flash | - | 1.00 | 1.00 | 1.00 | 1.00 | 1.00 | 1.00 | 1.00 | 1.00 |
| Qwen-Max | qwen-max | max | - | 1.00 | 1.00 | 1.00 | 1.00 | 1.00 | 1.00 | 1.00 | 1.00 |
| Qwen3-Max | qwen-max | 3-max | - | 1.00 | 1.00 | 1.00 | 1.00 | 1.00 | 1.00 | 1.00 | 1.00 |

| | | | | | | | | | | | |
|---|---|---|---|---|---|---|---|---|---|---|---|
| Qwen3.7-Max | qwen-max | 3.7-max | - | 1.00 | 1.00 | 1.00 | 1.00 | 1.00 | 1.00 | 1.00 | 1.00 |

**Table S3.** Mean sensitivity across conversation depths (therapy-transcript mean).

| Model | Family | Version | Params | d0 | d1 | d5 | d10 | d25 | d50 | d100 | d200 |
|---|---|---|---|---|---|---|---|---|---|---|---|
| Qwen 7B | qwen | 1.x | 7B | 0.780 | 0.786 | 0.644 | 0.641 | 0.598 | 0.555 | 0.556 | 0.338 |
| Qwen 14B | qwen | 1.x | 14B | 0.865 | 0.944 | 0.933 | 0.873 | 0.733 | 0.697 | 0.780 | 0.670 |
| Qwen 2 0.5B | qwen | 2 | 0.5B | 0.140 | 0.053 | 0.016 | 0.051 | 0.031 | 0.325 | 0.191 | 0.314 |
| Qwen 2 1.5B | qwen | 2 | 1.5B | 0.680 | 0.788 | 0.851 | 0.689 | 0.734 | 0.744 | 0.772 | 1.00 |
| Qwen 2 7B | qwen | 2 | 7B | 0.940 | 0.939 | 0.891 | 0.934 | 0.911 | 0.850 | 0.834 | 0.829 |
| Qwen 2.5 0.5B | qwen | 2.5 | 0.5B | 0.000 | 0.000 | 0.000 | 0.000 | 0.000 | 0.000 | 0.000 | 0.000 |
| Qwen 2.5 1.5B | qwen | 2.5 | 1.5B | 0.235 | 0.173 | 0.128 | 0.199 | 0.260 | 0.369 | 0.662 | 0.678 |
| Qwen 2.5 3B | qwen | 2.5 | 3B | 0.860 | 0.866 | 0.391 | 0.484 | 0.392 | 0.464 | 0.382 | 0.310 |
| Qwen 2.5 7B | qwen | 2.5 | 7B | 0.970 | 0.961 | 0.904 | 0.898 | 0.838 | 0.764 | 0.764 | 0.805 |
| Qwen 2.5 14B | qwen | 2.5 | 14B | 0.905 | 0.940 | 0.947 | 0.952 | 0.920 | 0.892 | 0.863 | 0.836 |
| Qwen 2 57B-A14B | qwen | 2 | 57B-A14B | 0.985 | 0.973 | 0.897 | 0.895 | 0.806 | 0.817 | 0.660 | 0.613 |
| Qwen 2.5 32B | qwen | 2.5 | 32B | 0.965 | 0.977 | 0.953 | 0.962 | 0.930 | 0.882 | 0.821 | 0.810 |
| Qwen 3 1.7B | qwen | 3 | 1.7B | 0.675 | 0.745 | 0.771 | 0.757 | 0.481 | 0.419 | 0.274 | 0.212 |
| Qwen 3 4B | qwen | 3 | 4B | 0.935 | 0.964 | 0.908 | 0.927 | 0.928 | 0.899 | 0.846 | 0.756 |
| Qwen 3 8B | qwen | 3 | 8B | 0.785 | 0.916 | 0.905 | 0.943 | 0.961 | 0.925 | 0.878 | 0.810 |
| Qwen 3 14B | qwen | 3 | 14B | 0.945 | 0.946 | 0.916 | 0.930 | 0.943 | 0.884 | 0.852 | 0.792 |
| Qwen 3 32B | qwen | 3 | 32B | 0.870 | 0.799 | 0.727 | 0.754 | 0.775 | 0.772 | 0.755 | 0.755 |
| Qwen 3.5 2B | qwen | 3.5 | 2B | 0.750 | 0.908 | 0.846 | 0.857 | 0.832 | 0.877 | 0.947 | 0.980 |
| Qwen 3.5 4B | qwen | 3.5 | 4B | 0.740 | 0.748 | 0.778 | 0.779 | 0.781 | 0.760 | 0.757 | 0.742 |
| Qwen 3.5 9B | qwen | 3.5 | 9B | 0.950 | 0.940 | 0.882 | 0.877 | 0.852 | 0.847 | 0.825 | 0.817 |
| Qwen 3.5 27B | qwen | 3.5 | 27B | 0.940 | 0.938 | 0.909 | 0.891 | 0.884 | 0.893 | 0.852 | 0.826 |
| Qwen 3.5 35B-a3b | qwen | 3.5 | 35B-A3B | 0.870 | 0.903 | 0.886 | 0.864 | 0.859 | 0.856 | 0.802 | 0.772 |
| Gemma 2 2B | gemma | 2 | 2B | 0.395 | 0.375 | 0.432 | 0.543 | 0.689 | 0.539 | 0.533 | 0.000 |

| | | | | | | | | | | | |
|---|---|---|---|---|---|---|---|---|---|---|---|
| Gemma 2 9B | gemma | 2 | 9B | 0.805 | 0.867 | 0.892 | 0.902 | 0.902 | 0.831 | 0.779 | 0.000 |
| Gemma 2 27B | gemma | 2 | 27B | 0.905 | 0.980 | 0.978 | 0.979 | 0.961 | 0.904 | 0.725 | 0.000 |
| Gemma 3 0.27B | gemma | 3 | 0.27B | 0.875 | 0.345 | 0.155 | 0.010 | 0.000 | 0.001 | 0.000 | 0.000 |
| Gemma 3 1B | gemma | 3 | 1B | 0.635 | 0.885 | 0.967 | 0.852 | 0.850 | 0.771 | 0.819 | 0.129 |
| Gemma 3 4B | gemma | 3 | 4B | 0.865 | 0.832 | 0.800 | 0.913 | 0.927 | 0.967 | 0.974 | 0.979 |
| Gemma 3 12B | gemma | 3 | 12B | 0.970 | 0.983 | 0.981 | 0.992 | 0.975 | 0.960 | 0.900 | 0.854 |
| Gemma 3 27B | gemma | 3 | 27B | 0.955 | 0.979 | 0.979 | 0.983 | 0.984 | 0.988 | 0.990 | 0.936 |
| Gemma 4 E4B | gemma | 4 | E4B | 0.975 | 0.966 | 0.599 | 0.772 | 0.971 | 0.972 | 0.981 | 0.964 |
| Gemma 4 26B | gemma | 4 | 26B-A4B | 0.965 | 0.955 | 0.935 | 0.932 | 0.925 | 0.944 | 0.929 | 0.892 |
| Gemma 4 31B | gemma | 4 | 31B | 0.965 | 0.972 | 0.966 | 0.943 | 0.933 | 0.923 | 0.910 | 0.905 |
| LLaMA 2 7B | llama | 2 | 7B | 0.535 | 0.522 | 0.446 | 0.413 | 0.701 | 0.518 | 0.000 | 0.000 |
| LLaMA 2 13B | llama | 2 | 13B | 0.000 | 0.053 | 0.152 | 0.186 | 0.091 | 0.002 | 0.000 | 0.000 |
| LLaMA 3.0 8B | llama | 3.0 | 8B | 0.845 | 0.903 | 0.942 | 0.924 | 0.774 | 0.496 | 0.302 | 0.000 |
| LLaMA 3.1 8B | llama | 3.1 | 8B | 0.860 | 0.954 | 0.993 | 0.988 | 0.991 | 0.968 | 0.949 | 0.855 |
| LLaMA 3.2 1B | llama | 3.2 | 1B | 0.050 | 0.084 | 0.010 | 0.000 | 0.011 | 0.000 | 0.001 | 0.000 |
| LLaMA 3.2 3B | llama | 3.2 | 3B | 0.660 | 0.848 | 0.809 | 0.879 | 0.839 | 0.796 | 0.859 | 0.774 |
| Llama 4 Scout 17B-16E | llama | 4 | 17B-16E | 0.965 | 0.977 | 0.973 | 0.989 | 0.988 | 0.972 | 0.951 | 0.899 |
| GPT-3.5-turbo | gpt | 3.5 | - | 0.820 | 0.888 | 0.925 | 0.933 | 0.928 | 0.917 | 0.887 | 0.782 |
| GPT-4o | gpt | 4o | - | 0.955 | 0.936 | 0.938 | 0.947 | 0.956 | 0.964 | 0.960 | 0.943 |
| GPT-5.4 | gpt | 5.4 | - | 0.930 | 0.935 | 0.927 | 0.925 | 0.926 | 0.918 | 0.891 | 0.881 |
| Gemini 2.5 Flash | gemini | 2.5-flash | - | 0.925 | 0.937 | 0.908 | 0.905 | 0.905 | 0.911 | 0.894 | 0.889 |
| Gemini 3 Flash-preview | gemini | 3-flash-preview | - | 0.920 | 0.951 | 0.960 | 0.963 | 0.937 | 0.935 | 0.938 | 0.933 |
| Gemini 3.5 Flash | gemini | 3.5-flash | - | 0.925 | 0.949 | 0.961 | 0.978 | 0.968 | 0.976 | 0.962 | 0.951 |
| Qwen-Max | qwen-max | max | - | 0.960 | 0.973 | 0.961 | 0.971 | 0.956 | 0.963 | 0.952 | 0.912 |
| Qwen3-Max | qwen-max | 3-max | - | 0.960 | 0.976 | 0.975 | 0.972 | 0.968 | 0.964 | 0.959 | 0.943 |

| | | | | | | | | | | |
|---|---|---|---|---|---|---|---|---|---|---|
| Qwen3.7-Max | qwen-max | 3.7-max | - | 0.960 | 0.964 | 0.953 | 0.954 | 0.944 | 0.950 | 0.941 | 0.914 |

**Table S4.** Mean specificity across conversation depths (therapy-transcript mean).

| Model | Family | Version | Params | d0 | d1 | d5 | d10 | d25 | d50 | d100 | d200 |
|---|---|---|---|---|---|---|---|---|---|---|---|
| Qwen 7B | qwen | 1.x | 7B | 0.935 | 0.872 | 0.974 | 0.964 | 0.855 | 0.796 | 0.707 | 0.992 |
| Qwen 14B | qwen | 1.x | 14B | 0.895 | 0.707 | 0.592 | 0.636 | 0.701 | 0.776 | 0.556 | 0.543 |
| Qwen 2 0.5B | qwen | 2 | 0.5B | 0.860 | 0.530 | 0.794 | 0.939 | 0.921 | 0.660 | 0.472 | 0.086 |
| Qwen 2 1.5B | qwen | 2 | 1.5B | 0.565 | 0.579 | 0.502 | 0.583 | 0.383 | 0.389 | 0.367 | 0.000 |
| Qwen 2 7B | qwen | 2 | 7B | 0.950 | 0.913 | 0.910 | 0.705 | 0.596 | 0.694 | 0.549 | 0.588 |
| Qwen 2.5 0.5B | qwen | 2.5 | 0.5B | 0.000 | 0.678 | 1.00 | 1.00 | 0.963 | 0.988 | 1.00 | 0.863 |
| Qwen 2.5 1.5B | qwen | 2.5 | 1.5B | 1.00 | 0.994 | 1.00 | 1.00 | 0.995 | 0.793 | 0.548 | 0.365 |
| Qwen 2.5 3B | qwen | 2.5 | 3B | 0.900 | 0.822 | 0.940 | 0.898 | 0.812 | 0.801 | 0.797 | 0.800 |
| Qwen 2.5 7B | qwen | 2.5 | 7B | 0.895 | 0.850 | 0.880 | 0.749 | 0.788 | 0.758 | 0.747 | 0.606 |
| Qwen 2.5 14B | qwen | 2.5 | 14B | 0.960 | 0.955 | 0.961 | 0.970 | 0.960 | 0.895 | 0.889 | 0.801 |
| Qwen 2 57B-A14B | qwen | 2 | 57B-A14B | 0.875 | 0.719 | 0.876 | 0.765 | 0.742 | 0.733 | 0.767 | 0.783 |
| Qwen 2.5 32B | qwen | 2.5 | 32B | 0.995 | 0.976 | 0.983 | 0.980 | 0.986 | 0.993 | 1.00 | 1.00 |
| Qwen 3 1.7B | qwen | 3 | 1.7B | 0.945 | 0.950 | 0.857 | 0.794 | 0.907 | 0.966 | 0.987 | 0.994 |
| Qwen 3 4B | qwen | 3 | 4B | 0.950 | 0.816 | 0.809 | 0.807 | 0.656 | 0.678 | 0.746 | 0.795 |
| Qwen 3 8B | qwen | 3 | 8B | 1.00 | 0.948 | 0.892 | 0.762 | 0.731 | 0.829 | 0.932 | 0.979 |
| Qwen 3 14B | qwen | 3 | 14B | 0.995 | 0.970 | 0.979 | 0.964 | 0.832 | 0.939 | 0.872 | 0.963 |
| Qwen 3 32B | qwen | 3 | 32B | 1.00 | 1.00 | 1.00 | 1.00 | 1.00 | 1.00 | 1.00 | 0.993 |
| Qwen 3.5 2B | qwen | 3.5 | 2B | 0.940 | 0.698 | 0.730 | 0.714 | 0.542 | 0.338 | 0.150 | 0.018 |
| Qwen 3.5 4B | qwen | 3.5 | 4B | 0.990 | 1.00 | 0.990 | 0.977 | 0.962 | 0.966 | 0.929 | 0.910 |
| Qwen 3.5 9B | qwen | 3.5 | 9B | 0.985 | 0.985 | 0.989 | 0.973 | 0.988 | 0.958 | 0.979 | 0.924 |
| Qwen 3.5 27B | qwen | 3.5 | 27B | 1.00 | 1.00 | 1.00 | 1.00 | 1.00 | 1.00 | 1.00 | 1.00 |
| Qwen 3.5 35B-a3b | qwen | 3.5 | 35B-A3B | 0.995 | 0.986 | 0.991 | 0.990 | 0.973 | 0.969 | 0.994 | 0.965 |
| Gemma 2 2B | gemma | 2 | 2B | 0.985 | 0.955 | 0.954 | 0.949 | 0.831 | 0.990 | 0.471 | 0.000 |

| | | | | | | | | | | | |
|---|---|---|---|---|---|---|---|---|---|---|---|
| Gemma 2 9B | gemma | 2 | 9B | 0.965 | 0.948 | 0.874 | 0.802 | 0.592 | 0.703 | 0.713 | 0.000 |
| Gemma 2 27B | gemma | 2 | 27B | 0.955 | 0.819 | 0.728 | 0.762 | 0.584 | 0.638 | 0.723 | 0.000 |
| Gemma 3 0.27B | gemma | 3 | 0.27B | 0.000 | 0.000 | 0.000 | 0.000 | 0.000 | 0.000 | 0.000 | 0.000 |
| Gemma 3 1B | gemma | 3 | 1B | 0.140 | 0.119 | 0.001 | 0.000 | 0.000 | 0.000 | 0.000 | 0.000 |
| Gemma 3 4B | gemma | 3 | 4B | 0.930 | 0.886 | 0.813 | 0.381 | 0.306 | 0.144 | 0.003 | 0.004 |
| Gemma 3 12B | gemma | 3 | 12B | 0.960 | 0.924 | 0.854 | 0.709 | 0.590 | 0.617 | 0.682 | 0.662 |
| Gemma 3 27B | gemma | 3 | 27B | 0.990 | 0.972 | 0.954 | 0.946 | 0.802 | 0.618 | 0.598 | 0.725 |
| Gemma 4 E4B | gemma | 4 | E4B | 0.900 | 0.923 | 0.577 | 0.705 | 0.817 | 0.815 | 0.784 | 0.674 |
| Gemma 4 26B | gemma | 4 | 26B-A4B | 0.990 | 0.982 | 0.980 | 0.963 | 0.966 | 0.961 | 0.971 | 0.969 |
| Gemma 4 31B | gemma | 4 | 31B | 1.00 | 1.00 | 0.991 | 0.994 | 0.994 | 0.989 | 0.991 | 0.990 |
| LLaMA 2 7B | llama | 2 | 7B | 0.110 | 0.062 | 0.308 | 0.227 | 0.201 | 0.042 | 0.000 | 0.000 |
| LLaMA 2 13B | llama | 2 | 13B | 0.000 | 0.031 | 0.340 | 0.207 | 0.204 | 0.000 | 0.000 | 0.000 |
| LLaMA 3.0 8B | llama | 3.0 | 8B | 0.005 | 0.196 | 0.403 | 0.347 | 0.354 | 0.576 | 0.470 | 0.000 |
| LLaMA 3.1 8B | llama | 3.1 | 8B | 0.935 | 0.768 | 0.456 | 0.376 | 0.303 | 0.436 | 0.443 | 0.655 |
| LLaMA 3.2 1B | llama | 3.2 | 1B | 0.030 | 0.154 | 0.000 | 0.000 | 0.000 | 0.000 | 0.000 | 0.000 |
| LLaMA 3.2 3B | llama | 3.2 | 3B | 0.950 | 0.767 | 0.824 | 0.730 | 0.603 | 0.589 | 0.525 | 0.599 |
| Llama 4 Scout 17B-16E | llama | 4 | 17B-16E | 0.990 | 0.913 | 0.878 | 0.802 | 0.614 | 0.537 | 0.607 | 0.642 |
| GPT-3.5-turbo | gpt | 3.5 | - | 0.990 | 0.975 | 0.954 | 0.922 | 0.705 | 0.749 | 0.748 | 0.797 |
| GPT-4o | gpt | 4o | - | 1.00 | 1.00 | 1.00 | 1.00 | 0.994 | 0.989 | 0.988 | 0.936 |
| GPT-5.4 | gpt | 5.4 | - | 1.00 | 1.00 | 1.00 | 1.00 | 1.00 | 1.00 | 1.00 | 1.00 |
| Gemini 2.5 Flash | gemini | 2.5-flash | - | 1.00 | 0.994 | 1.00 | 0.990 | 0.979 | 0.985 | 0.969 | 0.960 |
| Gemini 3 Flash-preview | gemini | 3-flash-preview | - | 1.00 | 1.00 | 0.994 | 0.991 | 0.990 | 0.989 | 0.985 | 0.976 |
| Gemini 3.5 Flash | gemini | 3.5-flash | - | 1.00 | 1.00 | 0.995 | 0.989 | 0.986 | 0.980 | 0.965 | 0.965 |
| Qwen-Max | qwen-max | max | - | 1.00 | 0.994 | 0.986 | 0.982 | 0.966 | 0.943 | 0.954 | 0.933 |
| Qwen3-Max | qwen-max | 3-max | - | 1.00 | 1.00 | 0.983 | 0.984 | 0.943 | 0.938 | 0.942 | 0.874 |

| | | | | | | | | | | | |
|---|---|---|---|---|---|---|---|---|---|---|---|
| Qwen3.7-Max | qwen-max | 3.7-max | - | 1.00 | 1.00 | 1.00 | 0.995 | 0.989 | 0.985 | 0.987 | 0.982 |

**Table S5.** Mean F1 score across conversation depths (therapy-transcript mean).

| Model | Family | Version | Params | d0 | d1 | d5 | d10 | d25 | d50 | d100 | d200 |
|---|---|---|---|---|---|---|---|---|---|---|---|
| Qwen 7B | qwen | 1.x | 7B | 0.846 | 0.824 | 0.770 | 0.762 | 0.674 | 0.618 | 0.551 | 0.482 |
| Qwen 14B | qwen | 1.x | 14B | 0.878 | 0.860 | 0.808 | 0.793 | 0.727 | 0.732 | 0.707 | 0.588 |
| Qwen 2 0.5B | qwen | 2 | 0.5B | 0.219 | 0.064 | 0.027 | 0.082 | 0.049 | 0.277 | 0.188 | 0.253 |
| Qwen 2 1.5B | qwen | 2 | 1.5B | 0.643 | 0.714 | 0.725 | 0.635 | 0.607 | 0.621 | 0.630 | 0.666 |
| Qwen 2 7B | qwen | 2 | 7B | 0.945 | 0.927 | 0.900 | 0.844 | 0.799 | 0.802 | 0.725 | 0.757 |
| Qwen 2.5 0.5B | qwen | 2.5 | 0.5B | 0.000 | 0.000 | 0.000 | 0.000 | 0.000 | 0.000 | 0.000 | 0.000 |
| Qwen 2.5 1.5B | qwen | 2.5 | 1.5B | 0.381 | 0.272 | 0.225 | 0.321 | 0.389 | 0.366 | 0.555 | 0.501 |
| Qwen 2.5 3B | qwen | 2.5 | 3B | 0.878 | 0.850 | 0.493 | 0.554 | 0.432 | 0.527 | 0.389 | 0.308 |
| Qwen 2.5 7B | qwen | 2.5 | 7B | 0.935 | 0.912 | 0.893 | 0.841 | 0.827 | 0.773 | 0.764 | 0.743 |
| Qwen 2.5 14B | qwen | 2.5 | 14B | 0.931 | 0.947 | 0.954 | 0.961 | 0.939 | 0.897 | 0.880 | 0.837 |
| Qwen 2 57B-A14B | qwen | 2 | 57B-A14B | 0.934 | 0.869 | 0.889 | 0.851 | 0.790 | 0.796 | 0.671 | 0.658 |
| Qwen 2.5 32B | qwen | 2.5 | 32B | 0.980 | 0.977 | 0.967 | 0.971 | 0.957 | 0.933 | 0.898 | 0.891 |
| Qwen 3 1.7B | qwen | 3 | 1.7B | 0.780 | 0.828 | 0.804 | 0.772 | 0.599 | 0.571 | 0.419 | 0.322 |
| Qwen 3 4B | qwen | 3 | 4B | 0.942 | 0.899 | 0.866 | 0.875 | 0.824 | 0.821 | 0.820 | 0.774 |
| Qwen 3 8B | qwen | 3 | 8B | 0.880 | 0.931 | 0.900 | 0.868 | 0.865 | 0.884 | 0.902 | 0.881 |
| Qwen 3 14B | qwen | 3 | 14B | 0.969 | 0.958 | 0.945 | 0.946 | 0.897 | 0.909 | 0.864 | 0.862 |
| Qwen 3 32B | qwen | 3 | 32B | 0.930 | 0.887 | 0.839 | 0.859 | 0.870 | 0.870 | 0.858 | 0.854 |
| Qwen 3.5 2B | qwen | 3.5 | 2B | 0.829 | 0.825 | 0.800 | 0.801 | 0.723 | 0.696 | 0.680 | 0.662 |
| Qwen 3.5 4B | qwen | 3.5 | 4B | 0.846 | 0.852 | 0.869 | 0.862 | 0.859 | 0.847 | 0.826 | 0.810 |
| Qwen 3.5 9B | qwen | 3.5 | 9B | 0.967 | 0.962 | 0.931 | 0.921 | 0.914 | 0.897 | 0.891 | 0.862 |
| Qwen 3.5 27B | qwen | 3.5 | 27B | 0.969 | 0.968 | 0.952 | 0.942 | 0.938 | 0.943 | 0.919 | 0.903 |
| Qwen 3.5 35B-a3b | qwen | 3.5 | 35B-A3B | 0.928 | 0.942 | 0.935 | 0.922 | 0.911 | 0.907 | 0.887 | 0.852 |
| Gemma 2 2B | gemma | 2 | 2B | 0.560 | 0.502 | 0.560 | 0.660 | 0.746 | 0.685 | 0.469 | 0.000 |

| | | | | | | | | | | | |
|---|---|---|---|---|---|---|---|---|---|---|---|
| Gemma 2 9B | gemma | 2 | 9B | 0.875 | 0.904 | 0.884 | 0.860 | 0.788 | 0.790 | 0.760 | 0.000 |
| Gemma 2 27B | gemma | 2 | 27B | 0.928 | 0.908 | 0.870 | 0.884 | 0.819 | 0.808 | 0.724 | 0.000 |
| Gemma 3 0.27B | gemma | 3 | 0.27B | 0.609 | 0.253 | 0.130 | 0.010 | 0.000 | 0.001 | 0.000 | 0.000 |
| Gemma 3 1B | gemma | 3 | 1B | 0.509 | 0.639 | 0.652 | 0.587 | 0.581 | 0.537 | 0.580 | 0.112 |
| Gemma 3 4B | gemma | 3 | 4B | 0.894 | 0.859 | 0.811 | 0.731 | 0.715 | 0.688 | 0.656 | 0.658 |
| Gemma 3 12B | gemma | 3 | 12B | 0.965 | 0.955 | 0.923 | 0.871 | 0.819 | 0.823 | 0.822 | 0.778 |
| Gemma 3 27B | gemma | 3 | 27B | 0.972 | 0.976 | 0.967 | 0.965 | 0.903 | 0.842 | 0.838 | 0.855 |
| Gemma 4 E4B | gemma | 4 | E4B | 0.940 | 0.947 | 0.575 | 0.740 | 0.902 | 0.902 | 0.895 | 0.844 |
| Gemma 4 26B | gemma | 4 | 26B-A4B | 0.977 | 0.968 | 0.956 | 0.946 | 0.944 | 0.952 | 0.949 | 0.927 |
| Gemma 4 31B | gemma | 4 | 31B | 0.982 | 0.985 | 0.978 | 0.968 | 0.962 | 0.954 | 0.948 | 0.945 |
| LLaMA 2 7B | llama | 2 | 7B | 0.441 | 0.388 | 0.398 | 0.335 | 0.527 | 0.371 | 0.000 | 0.000 |
| LLaMA 2 13B | llama | 2 | 13B | 0.000 | 0.050 | 0.187 | 0.170 | 0.125 | 0.002 | 0.000 | 0.000 |
| LLaMA 3.0 8B | llama | 3.0 | 8B | 0.595 | 0.668 | 0.755 | 0.721 | 0.649 | 0.464 | 0.352 | 0.000 |
| LLaMA 3.1 8B | llama | 3.1 | 8B | 0.894 | 0.879 | 0.784 | 0.758 | 0.742 | 0.777 | 0.763 | 0.781 |
| LLaMA 3.2 1B | llama | 3.2 | 1B | 0.050 | 0.093 | 0.010 | 0.000 | 0.011 | 0.000 | 0.001 | 0.000 |
| LLaMA 3.2 3B | llama | 3.2 | 3B | 0.772 | 0.820 | 0.821 | 0.826 | 0.759 | 0.725 | 0.749 | 0.718 |
| Llama 4 Scout 17B-16E | llama | 4 | 17B-16E | 0.977 | 0.948 | 0.929 | 0.905 | 0.836 | 0.805 | 0.823 | 0.812 |
| GPT-3.5-turbo | gpt | 3.5 | - | 0.896 | 0.928 | 0.939 | 0.928 | 0.847 | 0.858 | 0.842 | 0.794 |
| GPT-4o | gpt | 4o | - | 0.977 | 0.967 | 0.967 | 0.973 | 0.974 | 0.976 | 0.974 | 0.941 |
| GPT-5.4 | gpt | 5.4 | - | 0.964 | 0.966 | 0.962 | 0.961 | 0.960 | 0.956 | 0.941 | 0.934 |
| Gemini 2.5 Flash | gemini | 2.5-flash | - | 0.961 | 0.964 | 0.949 | 0.945 | 0.940 | 0.946 | 0.928 | 0.921 |
| Gemini 3 Flash-preview | gemini | 3-flash-preview | - | 0.958 | 0.974 | 0.977 | 0.977 | 0.962 | 0.961 | 0.960 | 0.953 |
| Gemini 3.5 Flash | gemini | 3.5-flash | - | 0.961 | 0.973 | 0.978 | 0.983 | 0.977 | 0.978 | 0.963 | 0.957 |
| Qwen-Max | qwen-max | max | - | 0.980 | 0.983 | 0.973 | 0.976 | 0.961 | 0.954 | 0.953 | 0.921 |
| Qwen3-Max | qwen-max | 3-max | - | 0.980 | 0.986 | 0.979 | 0.978 | 0.956 | 0.952 | 0.951 | 0.913 |

| | | | | | | | | | | | |
|---|---|---|---|---|---|---|---|---|---|---|---|
| Qwen3.7-Max | qwen-max | 3.7-max | - | 0.980 | 0.981 | 0.975 | 0.974 | 0.966 | 0.967 | 0.963 | 0.946 |

**Table S6.** Model family versus clinician performance change across conversation depth

| Model family (N models) | Δ slope (LLM − clinician) | Δ 95% CI | z | q (BH-FDR) |
|---|---|---|---|---|
| Gemma (11) | -0.0374 | (-0.0507, -0.0242) | -5.54 | <0.001*** |
| LLaMA (7) | -0.0297 | (-0.0435, -0.0159) | -4.22 | <0.001*** |
| Qwen (22) | -0.0206 | (-0.0326, -0.0086) | -3.37 | 0.001** |
| GPT (3) | -0.0097 | (-0.0217, 0.0023) | -1.59 | 0.164 |
| Gemini Flash (3) | -0.0052 | (-0.0169, 0.0064) | -0.88 | 0.381 |
| Qwen-Max (3) | -0.0088 | (-0.0205, 0.0028) | -1.49 | 0.164 |

Note. Licensed-clinician reference slope = 0.0027 (-0.0089, 0.0143). Δ = LLM slope − clinician slope. * q < .05, ** q < .01, *** q < .001.

**Table S7.** Difference in sensitivity between therapy and other conversation types for both LLMs and clinicians

| | Mean Δ sensitivity | 95% CI | q (BH-FDR) |
|---|---|---|---|
| **Psychoeducation** | | | |
| Qwen (22) | -0.0890 | (-0.3808, 0.2027) | 0.952 |
| Gemma (11) | -0.0305 | (-0.5023, 0.4414) | 0.952 |
| LLaMA (7) | -0.0504 | (-0.6676, 0.5668) | 0.952 |
| GPT (3) | -0.0105 | (-0.1216, 0.1006) | 0.952 |
| Gemini Flash (3) | 0.0116 | (-0.0818, 0.1050) | 0.952 |
| Qwen-Max (3) | -0.0135 | (-0.1552, 0.1281) | 0.952 |
| Clinicians (8) | 0.0130 | (-0.1451, 0.1711) | 0.851 |
| **MSJ** | | | |
| Qwen (22) | -0.1577 | (-0.2609, -0.0544) | 0.05* |
| Gemma (11) | -0.2640 | (-0.7962, 0.2681) | 0.952 |
| LLaMA (7) | -0.0582 | (-0.5974, 0.4809) | 0.952 |
| GPT (3) | -0.0676 | (-0.4224, 0.2872) | 0.952 |
| Gemini Flash (3) | -0.0448 | (-0.3639, 0.2742) | 0.952 |
| Qwen-Max (3) | -0.1244 | (-0.4893, 0.2406) | 0.952 |
| Clinicians (8) | 0.0130 | (-0.1078, 0.1338) | 0.806 |
| **Lorem ipsum** | | | |
| Qwen (22) | 0.0601 | (-0.0450, 0.1652) | 0.952 |
| Gemma (11) | -0.0206 | (-0.1049, 0.0638) | 0.952 |
| LLaMA (7) | 0.0147 | (-0.5840, 0.6134) | 0.962 |
| GPT (3) | 0.0087 | (-0.1067, 0.1240) | 0.952 |
| Gemini Flash (3) | 0.0254 | (-0.0133, 0.0640) | 0.952 |
| Qwen-Max (3) | 0.0244 | (-0.0071, 0.0558) | 0.952 |
| Clinicians (8) | 0.0280 | (-0.1157, 0.1717) | 0.659 |

Note. Δ sensitivity = control − therapy. LLM rows: depth-averaged marginal sensitivity from separate therapy and control sensitivity LMMs (eight preregistered depths, equal weight); family-level z-test on independent estimates; BH-FDR q = 0.05 across 12 directional tests (six families × psychoeducation and MSJ). Clinician rows: depth-matched panel mean across eight depths.. q < .05, ** q < .01, *** q < .001.

**Table S8.** Extended-depth linear mixed-effect model analyses of F1 (12-model subset)

| | Coefficient | SE | 95% CI | P Value |
|---|---|---|---|---|
| **Continued F1 change (therapy transcripts, standard arm)** | | | | |
| Intercept | 0.9807 | 0.0106 | [0.9599, 1.0015] | <0.001*** |
| log2(turns+1) | -0.0109 | 0.0010 | [-0.0128, -0.0089] | <0.001*** |
| Random effect: model SD | 0.0292 | | | |
| **Conversation-type effects beyond depth (four transcript types, standard arm)** | | | | |
| Intercept | 0.9804 | 0.0134 | [0.9541, 1.0066] | <0.001*** |
| log2(turns+1) | -0.0110 | 0.0008 | [-0.0125, -0.0094] | <0.001*** |
| Lorem Ipsum (vs therapy) | 0.0344 | 0.0074 | [0.0199, 0.0490] | <0.001*** |
| Psychoeducation (vs therapy) | 0.0165 | 0.0074 | [0.0020, 0.0311] | 0.03* |
| MSJ (vs therapy) | -0.0689 | 0.0074 | [-0.0835, -0.0544] | <0.001*** |
| Random effect: model SD | 0.0398 | | | |

Note. Mixed models on the 12-model deep subset (standard arm): F1 ~ log2(turns+1) + (1|model) for therapy transcripts; F1 ~ log2(turns+1) + conversation type + (1|model) for four transcript types, with therapy as reference. Conversation type explained variance beyond depth alone (likelihood-ratio test vs depth-only model, $\chi^2(3)=187.1$, P<0.001***). Exact Wilcoxon signed-rank tests on therapy deep-segment F1 change (n=12 models): change from 200 to max reached depth vs zero, median ΔF1 -0.009, P=0.24; deep vs shallow loss, median ΔF1 +0.053, P=0.2. P values are two-sided. CI = confidence interval.

**Table S9.** Extended depth F1 and ΔF1 at 0, 10, 200, and maximum depth for both standard and re-anchoring experiments

| | F1 (0) | F1 (10) | F1 (200) | F1 (max) | ΔF1 (10) | ΔF1 (200) | ΔF1 (max) | Δ (reanchor − std, max) | 95% CI | P Value |
|---|---|---|---|---|---|---|---|---|---|---|
| **Therapy** | | | | | | | | | | |
| Qwen 3.5 35B — Standard (1500) | 0.93 | 0.93 | 0.79 | 0.8 | 0 | -0.14 | -0.13 | - | | |
| Qwen 3.5 35B — Re-anchoring (1500) | 0.95 | 0.97 | 0.94 | 0.91 | 0.02 | -0.01 | -0.04 | 0.11 | | |
| Gemma 4 31B — Standard (1500) | 0.98 | 0.95 | 0.92 | 0.91 | -0.03 | -0.06 | -0.07 | - | | |
| Gemma 4 31B — Re-anchoring (1500) | 0.97 | 0.98 | 0.97 | 0.96 | 0.01 | 0 | -0.01 | 0.05 | | |
| Qwen 2.5 Max — Standard (500) | 0.98 | 0.97 | 0.89 | 0.83 | -0.01 | -0.09 | -0.15 | - | | |
| Qwen 2.5 Max — Re-anchoring (500) | 0.98 | 0.98 | 0.98 | 0.97 | 0 | 0 | -0.01 | 0.14 | | |
| Qwen3 Max — Standard (1500) | 0.98 | 0.97 | 0.92 | 0.81 | -0.01 | -0.06 | -0.17 | - | | |
| Qwen3 Max — Re-anchoring (1500) | 0.98 | 0.99 | 0.98 | 0.97 | 0.01 | 0 | -0.01 | 0.16 | | |
| Qwen3.7 Max — Standard (1500) | 0.98 | 0.96 | 0.92 | 0.95 | -0.02 | -0.06 | -0.03 | - | | |
| Qwen3.7 Max — Re-anchoring (1500) | 0.97 | 0.98 | 0.97 | 0.97 | 0.01 | 0 | 0 | 0.02 | | |
| Gemini 2.5 Flash — Standard (1500) | 0.96 | 0.93 | 0.9 | 0.92 | -0.03 | -0.06 | -0.04 | - | | |
| Gemini 2.5 Flash — Re-anchoring (1500) | 0.98 | 0.95 | 0.95 | 0.96 | -0.03 | -0.03 | -0.02 | 0.04 | | |
| Gemini 3 Flash — Standard (1500) | 0.96 | 0.96 | 0.91 | 0.7 | 0 | -0.05 | -0.26 | - | | |
| Gemini 3 Flash — Re-anchoring (1500) | 0.98 | 0.99 | 0.98 | 0.98 | 0.01 | 0 | 0 | 0.28 | | |
| Gemini 3.5 Flash — Standard (1500) | 0.96 | 0.98 | 0.92 | 0.92 | 0.02 | -0.04 | -0.04 | - | | |
| Gemini 3.5 Flash — Re-anchoring (1500) | 0.96 | 0.98 | 0.98 | 0.98 | 0.02 | 0.02 | 0.02 | 0.06 | | |
| GPT-3.5-turbo — Standard (200) | 0.9 | 0.91 | 0.64 | 0.64 | 0.01 | -0.26 | -0.26 | - | | |
| GPT-3.5-turbo — Re-anchoring (200) | 0.9 | 0.93 | 0.86 | 0.86 | 0.03 | -0.04 | -0.04 | 0.22 | | |
| GPT-4o — Standard (1500) | 0.98 | 0.97 | 0.92 | 0.76 | -0.01 | -0.06 | -0.22 | - | | |
| GPT-4o — Re-anchoring (1500) | 0.98 | 0.98 | 0.98 | 0.98 | 0 | 0 | 0 | 0.22 | | |
| GPT-5.4 — Standard (1500) | 0.96 | 0.95 | 0.9 | 0.9 | -0.01 | -0.06 | -0.06 | - | | |
| GPT-5.4 — Re-anchoring (1500) | 0.96 | 0.97 | 0.96 | 0.98 | 0.01 | 0 | 0.02 | 0.08 | | |
| Llama 4 Scout 17B — Standard (1500) | 0.98 | 0.92 | 0.82 | 0.8 | -0.06 | -0.16 | -0.18 | - | | |
| Llama 4 Scout 17B — Re-anchoring (1500) | 0.98 | 0.95 | 0.95 | 0.94 | -0.03 | -0.03 | -0.04 | 0.14 | | |
| *Pooled signed-rank test (n=12)* | | | | | | | | *+0.12* | *[0.0673, 0.1798]* | *<0.001**** |
| **Psychoeducation** | | | | | | | | | | |

| | | | | | | | | | | |
|---|---|---|---|---|---|---|---|---|---|---|
| Qwen 3.5 35B — Standard (1500) | 0.93 | 0.84 | 0.89 | 0.88 | -0.09 | -0.04 | -0.05 | - | | |
| Qwen 3.5 35B — Re-anchoring (1500) | 0.95 | 0.91 | 0.91 | 0.9 | -0.04 | -0.04 | -0.05 | 0.02 | | |
| Gemma 4 31B — Standard (1500) | 0.98 | 0.98 | 0.94 | 0.98 | 0 | -0.04 | 0 | - | | |
| Gemma 4 31B — Re-anchoring (1500) | 0.97 | 0.98 | 0.98 | 0.97 | 0.01 | 0.01 | 0 | -0.01 | | |
| Qwen 2.5 Max — Standard (500) | 0.98 | 0.98 | 0.95 | 0.94 | 0 | -0.03 | -0.04 | - | | |
| Qwen 2.5 Max — Re-anchoring (500) | 0.98 | 0.99 | 0.98 | 0.98 | 0.01 | 0 | 0 | 0.04 | | |
| Qwen3 Max — Standard (1500) | 0.98 | 0.97 | 0.94 | 0.96 | -0.01 | -0.04 | -0.02 | - | | |
| Qwen3 Max — Re-anchoring (1500) | 0.98 | 0.98 | 0.99 | 0.98 | 0 | 0.01 | 0 | 0.02 | | |
| Qwen3.7 Max — Standard (1500) | 0.98 | 0.93 | 0.94 | 0.97 | -0.05 | -0.04 | -0.01 | - | | |
| Qwen3.7 Max — Re-anchoring (1500) | 0.97 | 0.96 | 0.96 | 0.97 | -0.01 | -0.01 | 0 | 0 | | |
| Gemini 2.5 Flash — Standard (1500) | 0.96 | 0.91 | 0.84 | 0.93 | -0.05 | -0.12 | -0.03 | - | | |
| Gemini 2.5 Flash — Re-anchoring (1500) | 0.98 | 0.95 | 0.94 | 0.97 | -0.03 | -0.04 | -0.01 | 0.04 | | |
| Gemini 3 Flash — Standard (1500) | 0.96 | 0.98 | 0.96 | 0.97 | 0.02 | 0 | 0.01 | - | | |
| Gemini 3 Flash — Re-anchoring (1500) | 0.98 | 0.98 | 0.98 | 0.97 | 0 | 0 | -0.01 | 0 | | |
| Gemini 3.5 Flash — Standard (1500) | 0.96 | 0.98 | 0.97 | 0.98 | 0.02 | 0.01 | 0.02 | - | | |
| Gemini 3.5 Flash — Re-anchoring (1500) | 0.96 | 0.98 | 0.98 | 0.98 | 0.02 | 0.02 | 0.02 | 0 | | |
| GPT-3.5-turbo — Standard (350) | 0.9 | 0.89 | 0.68 | 0.79 | -0.01 | -0.22 | -0.11 | - | | |
| GPT-3.5-turbo — Re-anchoring (350) | 0.9 | 0.93 | 0.88 | 0.86 | 0.03 | -0.02 | -0.04 | 0.07 | | |
| GPT-4o — Standard (1500) | 0.98 | 0.97 | 0.98 | 0.99 | -0.01 | 0 | 0.01 | - | | |
| GPT-4o — Re-anchoring (1500) | 0.98 | 0.97 | 0.97 | 0.97 | -0.01 | -0.01 | -0.01 | -0.02 | | |
| GPT-5.4 — Standard (1500) | 0.96 | 0.95 | 0.96 | 0.96 | -0.01 | 0 | 0 | - | | |
| GPT-5.4 — Re-anchoring (1500) | 0.96 | 0.95 | 0.96 | 0.97 | -0.01 | 0 | 0.01 | 0.01 | | |
| Llama 4 Scout 17B — Standard (1500) | 0.98 | 0.88 | 0.78 | 0.7 | -0.1 | -0.2 | -0.28 | - | | |
| Llama 4 Scout 17B — Re-anchoring (1500) | 0.98 | 0.96 | 0.96 | 0.93 | -0.02 | -0.02 | -0.05 | 0.23 | | |
| *Pooled signed-rank test (n=12)* | | | | | | | | *+0.013* | *[-0.0006, 0.0560]* | *0.05* |
| **Lorem Ipsum** | | | | | | | | | | |
| Qwen 3.5 35B — Standard (1500) | 0.93 | 0.92 | 0.93 | 0.92 | -0.01 | 0 | -0.01 | - | | |
| Qwen 3.5 35B — Re-anchoring (1500) | 0.95 | 0.92 | 0.97 | 0.95 | -0.03 | 0.02 | 0 | 0.03 | | |
| Gemma 4 31B — Standard (1500) | 0.98 | 0.98 | 0.96 | 0.95 | 0 | -0.02 | -0.03 | - | | |

| | | | | | | | | | | |
|---|---|---|---|---|---|---|---|---|---|---|
| Gemma 4 31B — Re-anchoring (1500) | 0.97 | 0.99 | 0.98 | 0.97 | 0.02 | 0.01 | 0 | 0.02 | | |
| Qwen 2.5 Max — Standard (500) | 0.98 | 0.98 | 0.92 | 0.94 | 0 | -0.06 | -0.04 | - | | |
| Qwen 2.5 Max — Re-anchoring (500) | 0.98 | 0.99 | 0.99 | 0.97 | 0.01 | 0.01 | -0.01 | 0.03 | | |
| Qwen3 Max — Standard (1500) | 0.98 | 0.97 | 0.96 | 0.93 | -0.01 | -0.02 | -0.05 | - | | |
| Qwen3 Max — Re-anchoring (1500) | 0.98 | 0.99 | 0.97 | 0.98 | 0.01 | -0.01 | 0 | 0.05 | | |
| Qwen3.7 Max — Standard (1500) | 0.98 | 0.98 | 0.98 | 0.98 | 0 | 0 | 0 | - | | |
| Qwen3.7 Max — Re-anchoring (1500) | 0.97 | 0.98 | 0.99 | 0.98 | 0.01 | 0.02 | 0.01 | 0 | | |
| Gemini 2.5 Flash — Standard (1500) | 0.96 | 0.97 | 0.96 | 0.96 | 0.01 | 0 | 0 | - | | |
| Gemini 2.5 Flash — Re-anchoring (1500) | 0.98 | 0.96 | 0.98 | 0.97 | -0.02 | 0 | -0.01 | 0.01 | | |
| Gemini 3 Flash — Standard (1500) | 0.96 | 0.98 | 0.97 | 0.98 | 0.02 | 0.01 | 0.02 | - | | |
| Gemini 3 Flash — Re-anchoring (1500) | 0.98 | 0.98 | 0.98 | 0.98 | 0 | 0 | 0 | 0 | | |
| Gemini 3.5 Flash — Standard (1500) | 0.96 | 0.98 | 0.96 | 0.98 | 0.02 | 0 | 0.02 | - | | |
| Gemini 3.5 Flash — Re-anchoring (1500) | 0.96 | 0.98 | 0.98 | 0.98 | 0.02 | 0.02 | 0.02 | 0 | | |
| GPT-3.5-turbo — Standard (200) | 0.9 | 0.94 | 0.84 | 0.84 | 0.04 | -0.06 | -0.06 | - | | |
| GPT-3.5-turbo — Re-anchoring (200) | 0.9 | 0.9 | 0.84 | 0.84 | 0 | -0.06 | -0.06 | 0 | | |
| GPT-4o — Standard (1500) | 0.98 | 0.98 | 0.98 | 0.96 | 0 | 0 | -0.02 | - | | |
| GPT-4o — Re-anchoring (1500) | 0.98 | 0.98 | 0.98 | 0.98 | 0 | 0 | 0 | 0.02 | | |
| GPT-5.4 — Standard (1500) | 0.96 | 0.97 | 0.98 | 0.96 | 0.01 | 0.02 | 0 | - | | |
| GPT-5.4 — Re-anchoring (1500) | 0.96 | 0.96 | 0.97 | 0.96 | 0 | 0.01 | 0 | 0 | | |
| Llama 4 Scout 17B — Standard (1500) | 0.98 | 0.91 | 0.82 | 0.85 | -0.07 | -0.16 | -0.13 | - | | |
| Llama 4 Scout 17B — Re-anchoring (1500) | 0.98 | 0.94 | 0.94 | 0.94 | -0.04 | -0.04 | -0.04 | 0.09 | | |
| *Pooled signed-rank test (n=12)* | | | | | | | | *+0.009* | *[0.0024, 0.0437]* | *0.005*** |
| **MSJ** | | | | | | | | | | |
| Qwen 3.5 35B — Standard (1500) | 0.93 | 0.89 | 0.63 | 0.55 | -0.04 | -0.3 | -0.38 | - | | |
| Qwen 3.5 35B — Re-anchoring (1500) | 0.95 | 0.89 | 0.68 | 0.62 | -0.06 | -0.27 | -0.33 | 0.07 | | |
| Gemma 4 31B — Standard (1500) | 0.98 | 0.98 | 0.71 | 0.63 | 0 | -0.27 | -0.35 | - | | |
| Gemma 4 31B — Re-anchoring (1500) | 0.97 | 0.97 | 0.92 | 0.92 | 0 | -0.05 | -0.05 | 0.29 | | |
| Qwen 2.5 Max — Standard (500) | 0.98 | 0.96 | 0.69 | 0.67 | -0.02 | -0.29 | -0.31 | - | | |
| Qwen 2.5 Max — Re-anchoring (500) | 0.98 | 0.98 | 0.77 | 0.78 | 0 | -0.21 | -0.2 | 0.11 | | |

| | | | | | | | | | | |
|---|---|---|---|---|---|---|---|---|---|---|
| Qwen3 Max — Standard (1500) | 0.98 | 0.97 | 0.61 | 0.49 | -0.01 | -0.37 | -0.49 | - | | |
| Qwen3 Max — Re-anchoring (1500) | 0.98 | 0.96 | 0.79 | 0.73 | -0.02 | -0.19 | -0.25 | 0.24 | | |
| Qwen3.7 Max — Standard (1500) | 0.98 | 0.97 | 0.8 | 0.76 | -0.01 | -0.18 | -0.22 | - | | |
| Qwen3.7 Max — Re-anchoring (1500) | 0.97 | 0.98 | 0.96 | 0.86 | 0.01 | -0.01 | -0.11 | 0.1 | | |
| Gemini 2.5 Flash — Standard (1500) | 0.96 | 0.96 | 0.74 | 0.77 | 0 | -0.22 | -0.19 | - | | |
| Gemini 2.5 Flash — Re-anchoring (1500) | 0.98 | 0.95 | 0.82 | 0.76 | -0.03 | -0.16 | -0.22 | -0.01 | | |
| Gemini 3 Flash — Standard (1500) | 0.96 | 0.98 | 0.76 | 0.78 | 0.02 | -0.2 | -0.18 | - | | |
| Gemini 3 Flash — Re-anchoring (1500) | 0.98 | 0.98 | 0.95 | 0.93 | 0 | -0.03 | -0.05 | 0.15 | | |
| Gemini 3.5 Flash — Standard (1500) | 0.96 | 0.99 | 0.94 | 0.97 | 0.03 | -0.02 | 0.01 | - | | |
| Gemini 3.5 Flash — Re-anchoring (1500) | 0.96 | 0.98 | 0.96 | 0.96 | 0.02 | 0 | 0 | -0.01 | | |
| GPT-3.5-turbo — Standard (350) | 0.9 | 0.93 | 0.61 | 0.61 | 0.03 | -0.29 | -0.29 | - | | |
| GPT-3.5-turbo — Re-anchoring (200) | 0.9 | 0.92 | 0.78 | 0.78 | 0.02 | -0.12 | -0.12 | 0.17 | | |
| GPT-4o — Standard (1500) | 0.98 | 0.98 | 0.81 | 0.85 | 0 | -0.17 | -0.13 | - | | |
| GPT-4o — Re-anchoring (1500) | 0.98 | 0.98 | 0.81 | 0.75 | 0 | -0.17 | -0.23 | -0.1 | | |
| GPT-5.4 — Standard (1500) | 0.96 | 0.95 | 0.94 | 0.96 | -0.01 | -0.02 | 0 | - | | |
| GPT-5.4 — Re-anchoring (1500) | 0.96 | 0.96 | 0.95 | 0.94 | 0 | -0.01 | -0.02 | -0.02 | | |
| Llama 4 Scout 17B — Standard (1500) | 0.98 | 0.96 | 0.79 | 0.75 | -0.02 | -0.19 | -0.23 | - | | |
| Llama 4 Scout 17B — Re-anchoring (1500) | 0.98 | 0.98 | 0.88 | 0.82 | 0 | -0.1 | -0.16 | 0.07 | | |
| *Pooled signed-rank test (n=12)* | | | | | | | | *+0.085* | *[0.0019, 0.1672]* | *0.04** |

**Table S10.** Mixed model on paired reanchor−standard F1 differences (12-model subset)

| | Coefficient | SE | 95% CI | P Value |
|---|---|---|---|---|
| Intercept | 0.0108 | 0.0072 | [-0.0032, 0.0249] | 0.13 |
| log2(turns+1) | 0.0062 | 0.0006 | [0.0050, 0.0073] | <0.001*** |
| Lorem Ipsum (vs therapy) | -0.0335 | 0.0053 | [-0.0439, -0.0231] | <0.001*** |
| Psychoeducation (vs therapy) | -0.0210 | 0.0053 | [-0.0314, -0.0106] | <0.001*** |
| MSJ (vs therapy) | 0.0015 | 0.0053 | [-0.0089, 0.0119] | 0.78 |
| Random effect: model SD | 0.0180 | | | |

Note. dF1 = F1(re-anchoring) − F1(standard). Mixed model: dF1 ~ log2(turns+1) + conversation type + (1|model); therapy reference. P values are two-sided. CI = confidence interval.

**Table S11.** Preregistered primary (H1–H4) and extension (EH1–EH3) hypotheses with inferential results, support verdicts, and backing-table pointers.

| Hypothesis | Tier | Prediction | Preregistered test | Analysis performed | Result | Support | Data | Deviation from preregistration |
|---|---|---|---|---|---|---|---|---|
| H1 | primary | LLM F1 degrades with conversation depth. | Per family: test log2(turns+1) coefficient in F1 ~ log2(turns+1) + log2(parameters) + version + (1\|conversation) + (1\|model_id); support if coefficient < 0 and p < .05 (two-tailed). | Same family-specific LMM on therapy transcripts; coefficient p-value from mixed-effects fit (Table 1). | Negative log2(turns+1) slopes in all six LLM families (β −0.035 to −0.003, all p<.001; Table 1) | supported | Table 1 | Eight preregistered open-weight models not evaluated (could not run on study hardware): Qwen 72B, Qwen 2 72B, Qwen 2.5 72B, Gemma 4 E2B, LLaMA 2 70B, LLaMA 3.0 70B, LLaMA 3.1 70B, LLaMA 3.2 11B-vision. Analytic sample 49 models vs prereg target 59. Anthropic Opus 3, Opus 4, Opus 4.7 omitted per preregistered budget clause (not a deviation). Added LLaMA Scout v4 to introduce SoTA LLaMA open-weifht model |
| H2a | primary | Larger open-weight models outperform smaller open-weight models. | Per open-weight family: test log2(parameters) coefficient > 0 and p < .05 in the family LMM. | Same specification on open-weight families in Table 1; omitted for frontier families without public parameter counts. | Positive log2(parameters) in all three open-weight families (Qwen β=0.099, Gemma β=0.097, LLaMA β=0.296; all p<.05; Table 1). Parameter scaling was not tested for frontier families (not in scope). | supported | Table 1 | Same roster deviation as H1. |
| H2b | primary | Newer model versions outperform older versions within a family. | Per family with multiple versions: version dummy coefficients vs earliest reference; support if later versions positive and p < .05. | Same version-contrast structure in Table 1 family LMMs. | Significant newer-version advantages in Gemma (v4), LLaMA (3.x), GPT (v4, v5), and Gemini Flash (v3, 3.x); Qwen version dummies ns (Table 1) | supported | Table 1 | Same roster deviation as H1. |
| H3 (clinician LMM) | primary | Clinician F1 does not degrade with conversation depth. | Clinician LMM F1 ~ log2(turns+1) + (1\|conversation) + (1\|clinician_id); support if log2(turns+1) coefficient is not significant (two-tailed). | Same clinician LMM on therapy transcripts; coefficient p-value from mixed-effects fit (Table 1). | Clinician depth slope β=0.003, 95% CI includes 0 (ns) | supported | Table 1 | None. |
| H3 (cross-arm) | primary | Clinicians degrade less than LLMs across conversation depth. | z = (β_LLM − β_clinician) / sqrt(SE²_LLM + SE²_clinician) for each LLM family/provider; BH-FDR q = 0.05 across seven comparisons. | Same z-test on therapy depth slopes; BH-FDR q = 0.05 across six LLM families (Table S6). | Significant (q<.05): Qwen, Gemma, LLaMA; not significant: GPT, Gemini Flash, Qwen-Max | supported | Table S6 | Same roster deviation as H1. |
| H4a | primary | LLM sensitivity shifts by control condition: psychoeducation > therapy; many-shot jailbreak | Per family and directional control (psychoeducation, MSJ): z-test comparing depth-averaged marginal sensitivity from control vs therapy LMMs; BH-FDR q = 0.05 across 14 tests (seven families × two directional | Per family and control: depth-averaged marginal sensitivity from separate therapy and control sensitivity LMMs; z-test on independent | Psychoed>therapy (predicted): significant correct direction — none; significant wrong direction — none; not significant — Qwen, Gemma, LLaMA, GPT, Gemini Flash, Qwen-Max. MSJ<therapy | mostly unsupported | Table S7 | Same roster deviation as H1. |

| | | | | | | | | |
|---|---|---|---|---|---|---|---|---|
| | | (MSJ) < therapy. | controls). Lorem ipsum exploratory. | estimates; BH-FDR q = 0.05 across 12 directional tests (six families × psychoed and MSJ; lorem uncorrected) (Table S7). | (predicted): significant correct direction — Qwen; significant wrong direction — none; not significant — Gemma, LLaMA, GPT, Gemini Flash, Qwen-Max | | | |
| H4b | primary | Clinician sensitivity is not affected by control condition. | Clinician sensitivity contrasts for control vs therapy contexts; non-significant contrasts interpreted as no detected shift (two-tailed; no correction across three control tests). | Panel depth-matched Δsensitivity averaged across eight depths; one-sample two-tailed t-test vs zero per control type (Table S7). | Psychoeducation: p=0.851; MSJ: p=0.806; Lorem ipsum: p=0.659 | supported | Table S7 | Same roster deviation as H1. |
| EH1a | extension | F1 continues to degrade beyond the 200-turn ceiling (therapy, standard arm). | F1 ~ log2(turns+1) + (1\|model_id) on therapy merged transcript, standard arm; support if log2(turns+1) coefficient < 0 and p < .05 (two-tailed). | Same LMM on the preregistered 12-model deep subset (Table S8). | β=-0.0109, 95% CI [-0.0128, -0.0089], p=<0.001*** | supported | Table S8 | None. |
| EH1b | extension | Deep-segment F1 change (max reached length − 200 turns) differs from zero. | Two-sided exact Wilcoxon signed-rank test of ΔF1_deep = F1(max reached) − F1(200) across models (n = 12). | Same Wilcoxon test; median ΔF1 and P reported in Table S8 footnote. | median ΔF1=-0.0093, p=0.24, n=12. Post-200 segment does not differ from zero; continued 0–1500 decline significant in Table S8. | unsupported | Table S8 (footnote) | None. |
| EH1c | extension | Deep degradation exceeds shallow degradation (200 → max vs 0 → 200). | Two-sided exact Wilcoxon signed-rank test on paired (ΔF1_deep − ΔF1_shallow) across models (n = 12). | Same Wilcoxon test; median ΔF1 and P reported in Table S8 footnote. | median ΔF1=+0.0530, p=0.204, n=12. Deep-segment loss does not exceed shallow (0→200) loss. | unsupported | Table S8 (footnote) | None. |
| EH2a | extension | Conversation type adds explanatory variance beyond depth alone. | Likelihood-ratio test: F1 ~ log2(turns+1) + conversation + (1\|model_id) vs F1 ~ log2(turns+1) + (1\|model_id); two-tailed α = 0.05. | Same LRT; $\chi^2(3)$ and P reported in Table S8 footnote. | $\chi^2(3)$=187.102, p=<0.001 | supported | Table S8 (footnote) | None. |
| EH2b | extension | Psychoeducation and lorem coefficients positive; MSJ coefficient | Wald tests on conversation fixed effects in F1 ~ log2(turns+1) + conversation + (1\|model_id); two-tailed α = 0.05. | Same four-type LMM on 12-model subset; fixed-effect coefficients and P in Table S8. | Lorem: β=0.0344, p=<0.001***; Psychoeducation: β=0.0165, p=0.03*; MSJ: β=-0.0689, p=<0.001*** | supported | Table S8 | None. |

| | | | | | | | | |
|---|---|---|---|---|---|---|---|---|
| | | negative (vs therapy reference). | | | | | | |
| EH3a | extension | Reanchoring recovers F1 at each model's maximum reached in-window length. | Two-sided exact Wilcoxon signed-rank test on paired ΔF1 = F1(reanchor) − F1(standard) across models at each model's maximum reached in-window length, evaluated across all four conversation types (no multiplicity correction). | Four Wilcoxon tests (one per conversation type; n = 12 models each at model-specific max both-arm depth). Median ΔF1, 95% CI, and P in Table S9 pooled rows. | Therapy: median ΔF1=+0.1187, p=<0.001; Psychoeducation: median ΔF1=+0.0133, p=0.052; Lorem Ipsum: median ΔF1=+0.0090, p=0.005; MSJ: median ΔF1=+0.0846, p=0.042 | mixed | Table S9 | None. |
| EH3b | extension | Reanchoring recovery grows with conversation length. | Wald test on log2(turns+1) in ΔF1 ~ log2(turns+1) + conversation + (1\|model_id); support if coefficient > 0 and p < .05 (two-tailed). | Same paired-difference LMM on 12-model subset (Table S10). | β=0.0062, 95% CI [0.0050, 0.0073], p=<0.001*** | supported | Table S10 | None. |
| EH3c | extension | Reanchoring recovery is attenuated under MSJ relative to therapy. | Wald test on MSJ conversation coefficient in the EH3b LMM; support if coefficient < 0 and p < .05 (two-tailed). | Same paired-difference LMM on 12-model subset (Table S10). | β=0.0015, 95% CI [-0.0089, 0.0119], p=0.78 | unsupported | Table S10 | None. |
| QC-IRR | procedural | Clinician calibration quality control on held-out 50-probe IRR set (group Fleiss κ ≥ 0.40; individual median pairwise κ ≥ 0.40 or exclude). | Fleiss κ on 50-probe calibration set (8 clinicians); exclude/replace raters below individual threshold. | Fleiss/pairwise κ computed (Figure S15). All 8 clinicians above thresholds (pairwise κ 0.55–0.88; minimum median pairwise κ = 0.66); no replacements. | All 8 clinicians retained; pairwise κ range 0.55–0.88; min median pairwise κ=0.66 | passed | Figure S15 | None. |
| Secondary | secondary | Component-level depth effects, cross-family heterogeneity, robustness checks, moderators, and bootstrap uncertainty | (1) Parallel family LMMs with sensitivity, specificity, accuracy, and parse success as outcomes (descriptive p-values). (2) Joint cross-family F1 LMM. (3) Three sensitivity analyses: categorical depth, conversation:depth random effect, drop | Deferred; not run. | Deferred | | — | None. |

| | | |
|---|---|---|
| | intervals (no directional confirmatory claims). | extreme parameter sizes. (4) Severity and tone moderators with depth interactions. (5) Parametric bootstrap 95% CIs (B=1000, seed 42) for Table 1 contrasts. |
